# Updated Estimates of the Global, Regional and National Burden, and Etiology of Diarrheal Diseases Transmissible via Food: A Systematic Review and Meta-Analytical Modelling Study for the World Health Organization

**DOI:** 10.64898/2026.01.26.26344508

**Authors:** Josh M Colston, Brecht Devleesschauwer, Thomas Flynn, Francesca Schiaffino, Adhvikaa Ambikapathi Revathi, Nasif Hossain, Yen Ting Chen, Danielle Meister, Ava Bagherian, Olivia Binggeli, Emily Zheng, Nhiya David, Elaine Scallan Walter, Martyn D Kirk, Carlotta Di Bari, Robin Lake, Arie H Havelaar, Sara M Pires, Lucy J Robertson, Yuki Minato, Shannon E. Majowicz, Margaret N Kosek

## Abstract

Diarrheal disease is caused by diverse species of viruses, bacteria and protozo that are transmitted from different sources, including from contaminated food. Sustaining progress in reducing diarrheal illnesses and deaths, including vaccines and food safety measures, may require interventions targeting specific pathogens. In 2015, the WHO’s published etiology-specific estimates by their Foodborne Disease Burden Epidemiology Reference Group (FERG) of the incidence and mortality of diarrheal diseases caused by 11 pathogens at global and regional scales in 2010. Since then, much new evidence has been published about the epidemiology of enteropathogens. This study aims to update estimates to the year 2021 with the addition of three pathogens (Rotavirus, *Cyclospora cayetanensis*, Enteroaggregative *E. coli* (EAEC)). For low- and middle-income countries (LMICs), we conducted a systematic review of publications that reported the prevalence of pathogens diagnosed in stool samples from asymptomatic subjects, community-detected and outpatient diarrhea cases, and those treated as inpatients. Hierarchical, mixed effects models were fitted to pathogen-specific prevalence data extracted from studies that met prespecified inclusion criteria, and population attributable fractions (PAF) were calculated from the model parameter estimates, adjusting for background asymptomatic transmission where appropriate. The PAFs were applied to previously estimated diarrhea incidence and mortality envelopes. A separate, parallel systematic review identified studies that estimated diarrhea incidence and mortality due to specific etiologies for high income countries (HICs) from surveillance data. Meta-analytical models were fitted to data extracted from these. Data from 324 studies published between 1990 and 2023 representing results from up to 540,000 samples were used in the meta-analysis. Globally the 14 pathogens were responsible for 2.2 billion diarrheal disease cases and 880,000 deaths in 2021, with the largest number of cases occurring in South-East Asia, the largest number of deaths in Sub-Saharan Africa, and Europe being the region with the lowest burden by both measures. We found the leading causes of diarrhea morbidity to be bacteria – *Shigella* (426.4 million cases in 2021), *Campylobacter* (291.4 million) and ETEC (259.7 million)) – as well as the protozoon *Giardia* (321.2 million), in contrast to previous studies that have ranked rotavirus and norovirus highest. Our estimates support a much higher morbidity burden for *Shigella* than previously estimated with an incidence rate for 2021 (5,400 per 100,000). This is due to its high PAF in outpatients aged ≥5 years. As causes of diarrhea mortality, our estimates rank rotavirus first (214,700 deaths in 2021), *Shigella* second (152,500), and *V. cholerae* (94,100) third, due to the latter’s large PAF in inpatients aged ≥5 years and high case fatality rate. Caution is urged when interpreting PAFs for pathogens that elicit prolonged residual shedding following resolution of symptoms (e.g., norovirus, *Campylobacter*, *Giardia*). These findings, derived from rigorous systematic review and statistical methodologies and the largest database yet compiled of pathogen detection rates, will serve as inputs for the WHO’s broader estimates of hazard-specific incidence and mortality from foodborne diseases and are made available to the research and policy-making communities to inform targeted strategies for global diarrheal disease control.

**Funding:** WHO EDTF_001; 1K01AI168493-01A1; K43TW012298; 5T32AI007046-48.

## Introduction

Diarrheal disease is a common illness worldwide, particularly in children under the age of 5 years [1]. The syndrome is characterized by frequent passage of loose or watery stools, and is almost always the result of infections of the gastrointestinal tract by pathogens transmitted via the fecal-oral route [2]. In most cases these episodes are mild and resolve without specific therapy [3,4]. However, severe cases of diarrhea may cause serious illness, chronic sequelae or death. Since the turn of the 21^st^ century, tremendous and rapid progress has been made in reducing global deaths from diarrheal disease. Recent estimates rank it as the fastest declining cause of mortality in children under 5 years, having gone from the second to the sixth leading such cause since 2000 [5]. This is largely attributable to improved nutritional status and two simple yet highly cost-effective interventions: oral rehydration therapy (ORT), which prevents severe diarrhea from causing death from dehydration when administered to infants; and the rotavirus vaccine, which confers partial immunity to severe disease from a leading viral etiological agent of acute gastroenteritis [6]. However, rotavirus is just one pathogen among the various species and strains of widely endemic viruses, bacteria and protozoa – most of them with recognized foodborne transmission - that count among the etiologies of diarrheal disease [7]. Furthermore, high-quality, large, cluster-randomized trials of low-cost household-level water, sanitation, and hygiene (WASH) solutions have found at best only very modest reductions in pediatric diarrheal incidence [8–11] and enteropathogen prevalence in their study populations [12,13]. Sustaining the fragile progress of recent years, particularly in the face of threats such as climate change and US disengagement from global health financing, will require tools that target particular pathogens such as new vaccines, improved livestock management practices, and interventions to improve food safety. These will need to be introduced while maintaining coverage of existing interventions, such as monitoring and improving case management with oral rehydration therapy, the use of antibiotics in cases where clinically indicated, and reinforcement of nutrition, particularly during the first two years of life [14].

In 2015, the World Health Organization (WHO) and its technical advisory Foodborne Disease Burden Epidemiology Reference Group (FERG, convened from 2007 - 2015) published etiology-specific estimates of the incidence and mortality of diarrheal diseases caused by nine pathogens at global and regional scales for the year 2010 [15]. Separate systematic reviews also estimated the incidence and mortality due to Shiga toxin-producing *Eschrichia coli* (STEC) [16] and *Vibrio cholerae* [17]. Together the eleven diarrheal diseases assessed caused an estimated 1.9 billion (95% uncertainty interval [UI] 1.4-2.8 billion) cases and 715,000 (95% UI 603,000-846,000) deaths worldwide in 2010, though the authors noted the scarcity of available data as a key limitation in establishing reliable estimates [18]. The intervening years have seen continuing improvements in diagnostic technologies, multiple large-scale studies that have expanded the volume of high-quality data from diverse settings, and improved statistical methods have been developed, all of which have advanced the potential to attribute diarrheal episodes and burden to specific enteropathogens. The WHO reconvened FERG for 2021-2025 and commissioned a decadal follow-up of the 2015 publication to update the estimates, refine them in a way that reflects these developments, and extend them to additional diarrhea-causing pathogens now recognized as being commonly transmitted by food [19]. This article reports the methods, findings, and interpretation of this update.

## Materials and Methods

This study aimed to estimate the annual morbidity and mortality burden attributable to each of 14 diarrhea-causing pathogens commonly transmissible via food in the 194 WHO Member States, for each year from 2000 to 2021, and for two age groups (<5 and ≥5 years). We replicated the previous FERG approach and adapted it to reflect changes in understanding of the syndrome’s epidemiology in intervening years, as well as the addition of several pathogens not previously included. In addition to the 9 pathogens included in the 2015 study*—*thermophilic *Campylobacter* spp., *Cryptosporidium* spp., *Entamoeba histolytica*, enteropathogenic (EPEC) and enterotoxigenic *E. coli* (ETEC), *Giardia* spp., norovirus, nontyphoidal *Salmonella* spp., and *Shigella* spp.—for this iteration we also integrated STEC and *V. cholerae* within the same methodology (they had previously been estimated from separate systematic reviews), as well as adding to the list *Cyclospora cayetanensis*, enteroaggregative *E. coli* (EAEC), and rotavirus, bringing the number of agents to 14.

We took the same approach as the previous FERG study, where we estimated incidence separately for high income (HICs) and low- and middle-income countries (LMICs) to reflect a general dichotomy in the way that causes of diarrhea are monitored in the two groups of countries. While health information systems in many HICs directly and routinely monitor, report pathogen-specific diarrheal disease incidence and mortality, in most LMICs, which tend to have a higher overall burden, such direct surveillance is not yet available, and indirect methods of estimation are necessary. We categorized countries’ income status based on World Bank income level for 2024 (with some ad hoc modifications) [20,21].

### Approach 1 - Indirect etiology attribution for LMICs

For this group of countries, pathogen-specific population attributable fractions (PAFs) were calculated using a meta-analytical approach and these proportions applied to estimates of total diarrheal disease incidence and mortality (diarrheal and diarrheal death “envelopes” respectively) to derive pathogen-specific burden indicator values. Diarrheal incidence envelopes were obtained from GBD 2021, and mortality envelopes from the WHO’s Global Health Estimates, each stratified by age, sex, country, and year, for the years 2000 to 2021 [22,23].

**Data sources:** We sourced aggregate estimates of the prevalence of each endemic enteric pathogen from published population-based studies identified through a systematic review of the scientific literature (PROSPERO protocol CRD42023427998) [19]. We performed a structured search on the Medline-PubMed, Web of Science and Embase databases for articles published in peer-reviewed journals between January 1^st^ 1990 and June 30^th^ 2023 that reported pathogen-specific stool sample detection rates from human populations. These were imported into the *Covidence* software [24] and screened for eligibility in stages according to prespecified criteria to exclude the following:

- Studies with no primary data reported (reviews, meta-analyses etc.).
- Studies of only *in vitro* procedures, non-human subjects, or environmental samples.
- Studies that did not test for more than one of the 14 pathogens.
- Studies with fewer than 12 consecutive months of surveillance (or that did not state the duration).
- Studies that enrolled fewer than 100 subjects with diarrhea.
- Studies that assessed diarrhea solely in special or non-representative populations (HIV+ subjects, travelers, transplant patients, etc.).
- Studies with inclusion criteria for enrolment that did not use a standard definition of diarrhea or that were biased towards particular pathogen species or taxa (e.g., suspected viral diarrhea).
- Studies that recruited only asymptomatic subjects or without regard to diarrhea status.
- Studies that did not exclude nosocomial, persistent or antibiotic-associated diarrhea cases.
- Case reports or reports on outbreaks, epidemics or seasonal surges.
- Studies with insufficient information to calculate prevalence.
- Studies for which full texts could not be obtained following reasonable effort.
- Conference or meeting abstracts, preprints, and retracted publications.

From each eligible study, numerators (total positive stool samples for each pathogen tested) and denominators (total samples analyzed) were extracted within reported age, syndrome, and site strata according to a pre-specified template [19]. For studies for which microdata were available, either publicly or with permission of the investigators, we aggregated these totals within predefined strata (see Colston *et al*. 2025) [19], otherwise they were extracted as reported in the article. Syndrome strata included, in incrasing order of severity; 1). Asymptomatic subjects; 2). Outpatient (and community detected) diarrhea cases and 3). Inpatient diarrhea cases or deaths. For articles in which ETEC detections were disaggregated by heat-labile (LT) and heat-stable (ST) enterotoxins and EPEC by typical and atypical genotypes, these were extracted and modelled separately due to differences in their pathogenicity profiles and then reaggregated to their respective pathotypes once their separate modelled estimates had been obtained. Other variables extracted for use in the analysis included diagnostic method used (molecular [PCR] compared to conventional methods – culture for bacteria, microscopy for protozoa, and immunoassays for viruses), pathogen strain (categories varied by species), country, and whether the rotavirus vaccine had been introduced in that country at the time the data collection was carried out (information taken from VIEW-hub by IVAC) [25] Because etiology of diarrheal disease mortality is rarely reported in the literature, we used PAFs for inpatient (hospitalized) diarrheal cases as a proxy for death, since these represent the most severe episodes that are likeliest to result in death. We used PAFs from less-severe diarrhea cases detected in outpatient and community-based studies to estimate pathogen-specific incidence at the community level in endemic settings.

**Statistical methods:** For each pathogen, we fitted a hierarchical, mixed effects model to the logit-transformed prevalence proportions and corresponding standard errors with random effects specified at the levels of WHO region and sub-region, country, study, and stratum. Other variables were included as fixed effects to adjust for potential confounding introduced due to heterogeneity in study design and methodology, including syndrome (outpatient case or inpatient case compared to asymptomatic), age group (<5 years, mixed age groups, compared to ≥5 years), diagnostic method (conventional, compared to molecular), rotavirus vaccine introduction status (introduced compared to not introduced), and, where relevant, pathogen strain (e.g. genotypes, genogroups, species, enterotoxins etc.). Models were fitted using the *escalc* function from the *metafor* R package [26]. Model equations are reported in **Supplementary file 2**. Whereas FERG’s 2015 study employed the so-called “detection as etiology” (DE) assumption to define etiology attribution, whereby the PAF is defined simply as the normalized, crude prevalence of a pathogen in diarrheal samples [15,27], it has since become a consensus that the DE assumption overestimates the true PAF, at least for PCR-based detection, due to asymptomatic carriage of enteropathogens [28,29]. Recognizing this development, where possible this update adopted an odds-ratio (OR) method, whereby the PAF is calculated as a transformation of the ratio of the odds of detection of the pathogen in diarrheal compared to asymptomatic stool samples [30]. This is based on the assumption that for agents that are truly pathogenic there will be a positive dose-response relationship at the population level between diarrheal symptom severity and the detection rate of that pathogen in stool samples, such that the OR for detection will be >1, and the PAF >0. Exponentiated coefficients for each of the syndrome categories estimated by each of the models were treated as the pathogen (*g*)-specific prevalence in asymptomatic subjects, (*P***_ga_**), outpatient (*P***_go_**), and inpatient (*P***_gi_**) diarrheal cases. Therefore, for symptomatic syndrome stratum *x*:

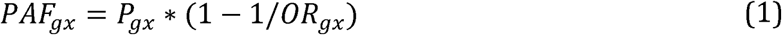

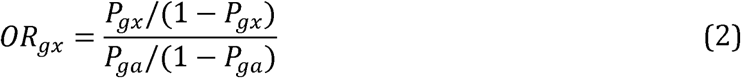

However, for some pathogens, this results in ORs of <1, leading to uninterpretable, negative PAFs. In such cases, a modified DE (mDE) approach was used, whereby the PAF was estimated as the average between the prevalence in symptomatic individuals (*P***_gx_** estimated from a model that excluded the asymptomatic syndrome category) and zero to reflect that these two extremes represent the maximum and minimum plausible range of the true value.

**Imputation:** After fitting the model to the available data, posterior predictive distributions were used to impute values for countries for which no data was available. The imputation process followed an algorithmic structure:

1. If data were available for other countries in the same subregion, the subregion-year-specific estimate was used.
2. If the subregion lacked data, but the broader region had data, the region-year-specific estimate was used.
3. If no data were available for the region, the global-year-specific estimate was applied.

### Approach 2 – Direct etiology-specific burden estimation for HICs

**Data sources:** For this group of countries, we sourced incidence and mortality data from published studies that estimated the national population-level incidence of illness caused by agents transmitted commonly through food (“burden of illness studies”) identified by updating a previously published scoping review [31]. The original review included studies published during January 1, 1995 to December 31, 2019 and was updated though December 31, 2023 for the purpose of informing these estimates, with methods described in detail in the previous publication [31]. Briefly, PubMed was queried using the terms: “Population surveillance and foodborne disease”, “Burden of illness and foodborne disease”, and “Community incidence and foodborne disease.” We first screened identified articles for relevance by reviewing titles and abstracts, then relevant publications underwent a full article review to verify that these were original estimates of the total number of illnesses. The references cited by selected articles were screened for other potentially eligible publications. We further restricted the search to articles presenting results for at least one of the 14 pathogens included as part of the WHO estimates and excluded articles conducted outside WHO subregion A countries (HICs) or with estimates limited to sub-populations (e.g., regional, specific age group) or a specific transmission route (e.g., food, water). Data on the estimated number or incidence of illnesses were extracted, along with the range of uncertainty in whatever metric they were presented (e.g. 95% uncertainty intervals, 90% credible intervals), for any of the 14 target pathogens (without distinguishing LT- from ST-ETEC). Data on the number or incidence of deaths was also extracted when available. EAEC and EPEC had insufficient data, so no estimated of the burden of those pathogens were extracted.

**Statistical analysis:** Given the relative scarcity of studies for HICs, we adopted a simplified modeling strategy for approach 2 compared to approach 1. For each combination of pathogen and burden indicator (incidence and mortality), four independent models were fitted to the log-transformed incidence and mortality values and corresponding standard errors, using the *escalc* function from the *metafor* package. These were defined by the inclusion or not of regional random effects, and of a global time trend. For each HIC, indicator estimates were taken from the output of the most appropriate model following a pre-defined algorithm that took into account the number of data points from that country, model convergence, and estimate uncertainty. Model equations are reported in supplementary file 2.

*Giardia* was assumed *a priori* by expert opinion not to be a cause of mortality in either HICs or LMICs [32] so only incidence attributable to that pathogen was estimated, and for *V. cholerae* incidence and mortality were only estimated for countries reporting endemic cholera transmission over the past 10 years [33], while countries deemed free from transmission were excluded from the imputation model and assigned a PAF of zero.

## Results

### Approach 1 - Indirect etiology attribution for LMICs

Approach 1 identified an initial retrieval set of almost 46,000 unique references (**Figure 1a**), of which 94.6% (43,365) were excluded based on a review of their titles and abstracts, and a further 4.4% (2,003) following review of their full texts, leaving just 1.0% (466) that reported on 414 eligible studies (“studies” are distinguished from “references” and “publications”, since some studies published findings on different pathogens over multiple publications). 127 of the remaining studies had data extracted but were not included in the model due to having taken place in HICs, resulting in 287 studies that contributed results to the estimates for LMICs. However, because numerous studies were conducted in multiple sites, often in different countries, the final dataset covered 332 locations in 61 different LMICs (**Figure 1b**).

**Figure 1:**
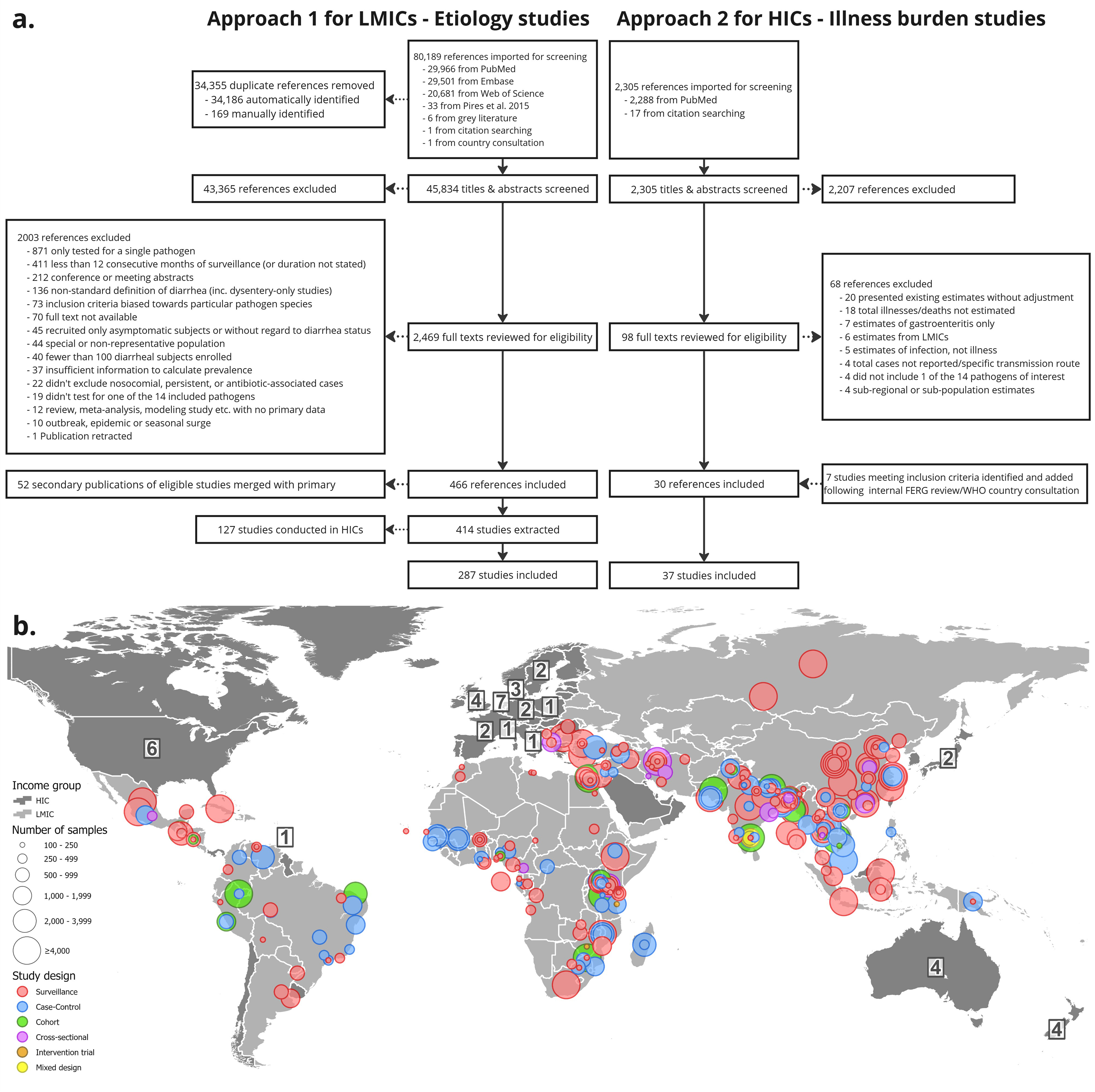
Results of the two systematic reviews: a). PRISMA-style flowchart of the numbers of studies included and excluded at each stage of the reviews for the two approaches; b). Locations of enrolment sites, study designs, and numbers of samples analyzed for eligible diarrhea etiology studies included in the model for approach 1. Numerical labels represent numbers per country of eligible burden of illness studies included in the by approach 2.

Rotavirus, *Shigella* and *Salmonella* were the three pathogens that were most frequently tested for (**Table 1**), with results for each available from more than 150 studies, and more than 50 countries. *C. cayetanensis* was by far the least represented pathogen in the overall database with just 18 studies reporting diagnostic results from 35 sites in 20 countries. EAEC was the pathogen with highest overall prevalence (18.0%) but was around twice as prevalent in asymptomatic individuals as in inpatient and four times as in outpatient diarrhea cases (40.4%, 20.1% and 8.6% respectively). This pattern was also seen with *Campylobacter*, but at lower rates of detection (8.8% overall, 19.0%, 10.1% and 5.8% respectively in asymptomatic subjects, inpatient and outpatient cases). Only 3 of the 14 pathogens strictly followed the hypothesized pattern of increasing prevalence of detection with increasing levels of symptom severity (rotavirus, *Salmonella* and *V. cholerae*), though most had higher positivity rates in inpatient than outpatient symptomatic cases. *Giardia,* by contrast, exhibited the exact inverse of the hypothesized pattern and starkly so, exhibiting decreasing prevalence with increasing symptom severity dropping from 27.6% in asymptomatic subjects to 12.4% in outpatient and 10.7% inpatient-attended diarrhea.

**Table 1:**
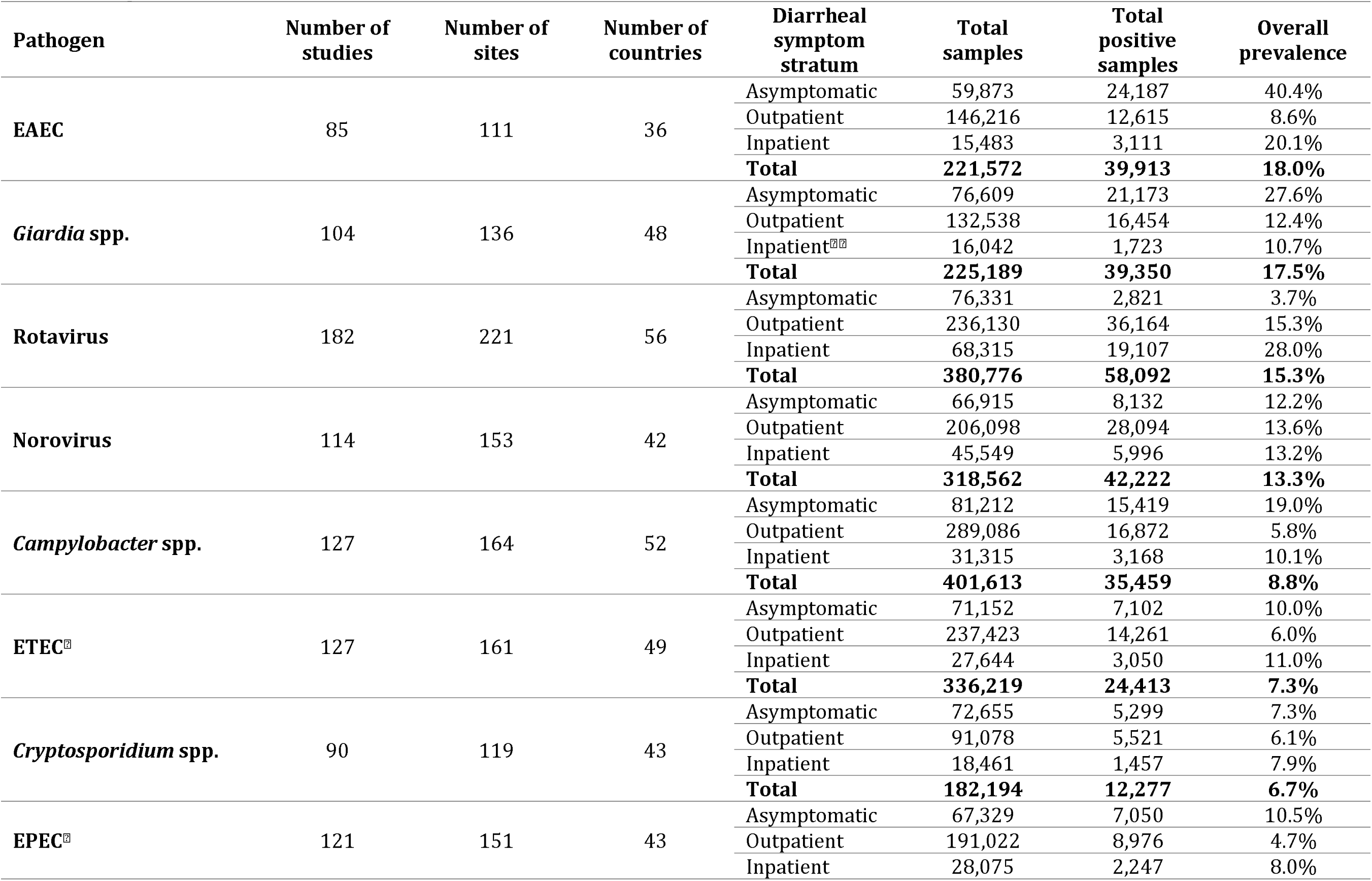
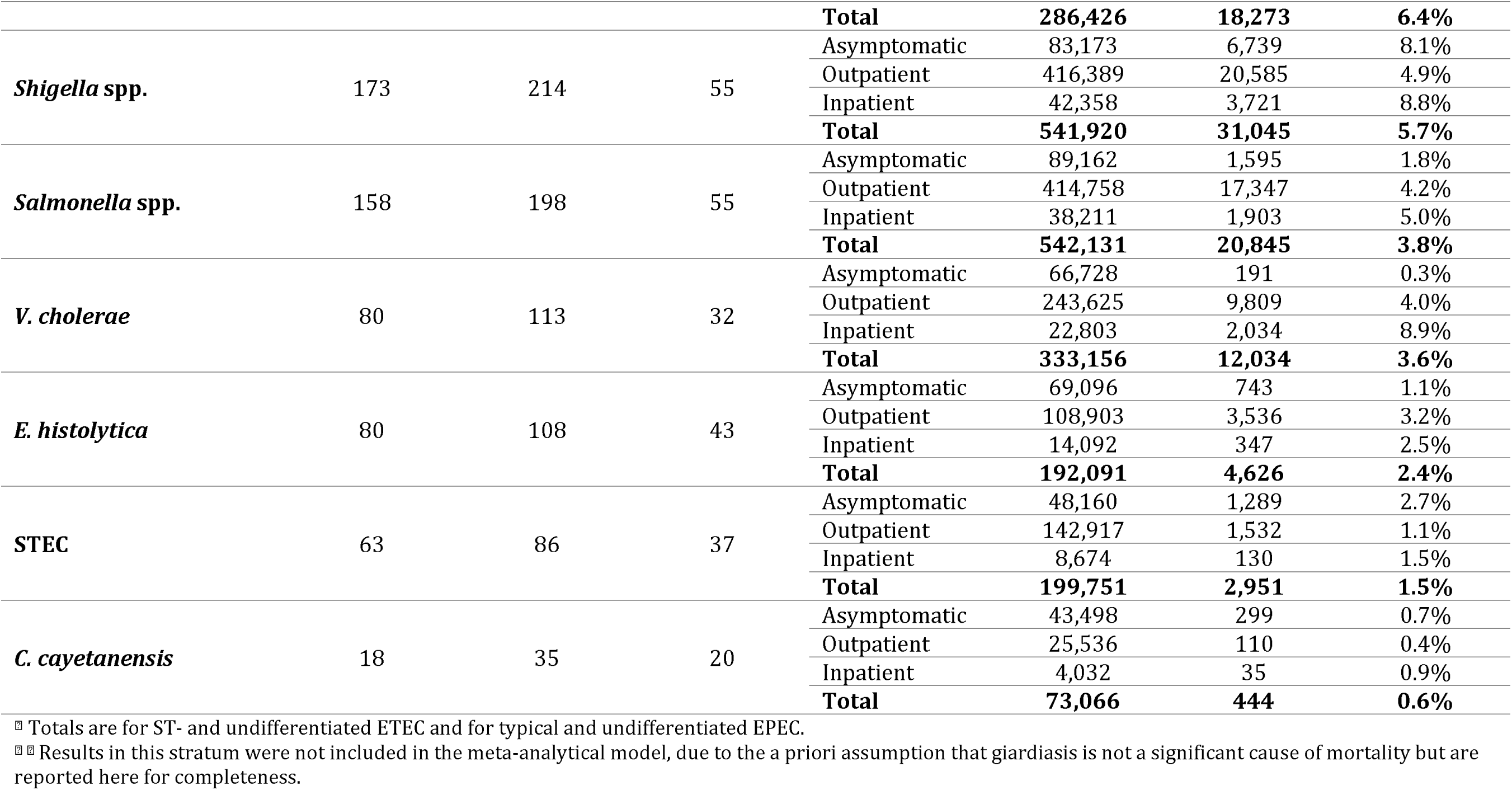
Numbers of studies, sites and countries for which diagnostic results were available for each of the pathogens using approach 1, ranked by total overall prevalence.

Upon adjusting for confounders in the multivariable logistic models, many of the apparent unexpected associations observed in the crude prevalences either reversed direction or amplified (**Figure 2**). For example, the crude relative prevalence of *Shigella* detection in outpatient diarrhea cases was 60.5% that of asymptomatic subjects, however, the adjusted odds ratio (aOR) comparing the same two categories indicated an almost 3-fold increase in the odds of *Shigella* positivity (aOR = 2.88 [2.29, 3.63]), while the equivalent association between inpatient cases and asymptomatic subjects went from just a 8.6% higher crude prevalence to 350% increased odds (3.50 [2.62, 4.62]). The equivalent changes for ST-ETEC were from 40% lower prevalence to 60% higher odds (1.61 [1.41, 1.85]) in outpatients, and from 10% higher prevalence to 70% higher odds (1.70 [1.39, 2.08]) in inpatients, while for EAEC, both associations reversed direction and attenuated towards the null (79% and 50% reduced prevalence aORs of 1.01 [0.89, 1.15] and 1.06 [0.88, 1.27] respectively in outpatients and inpatients compared to asymptomatic subjects). For 12 of the 15 pathogens (ST- and LT-ETEC having been analyzed separately at this stage), the models therefore yielded coefficient estimates that allowed for the use of the OR approach to calculating the PAFs, which adjusts for asymptomatic pathogen carriage (**Figure 2a**). For the remaining three pathogens - *Campylobacter*, *Giardia* and STEC - however, odds of detection in equivalent models (not shown) were lower in the symptomatic categories than in asymptomatic subjects, resulting in an apparent inverse association between infection and morbidity even upon adjustment, and necessitating a default to the mDE approach. Therefore, models for these three pathogens excluded results from asymptomatic subjects and instead treated inpatient cases as the referent and outpatient cases as the comparator categories (**Figure 2b**). Even with these modifications, aORs in outpatient diarrhea cases, relative to inpatient diarrhea cases, were small, statistically non-significant and in the opposite direction to the *a priori* hypothesis (aORs of 1.15 [0.90, 1.47], 1.33 [0.94, 1.90], and 1.01 [0.69, 1.46] respectively for *Campylobacter*, *Giardia* and STEC).

**Figure 2:**
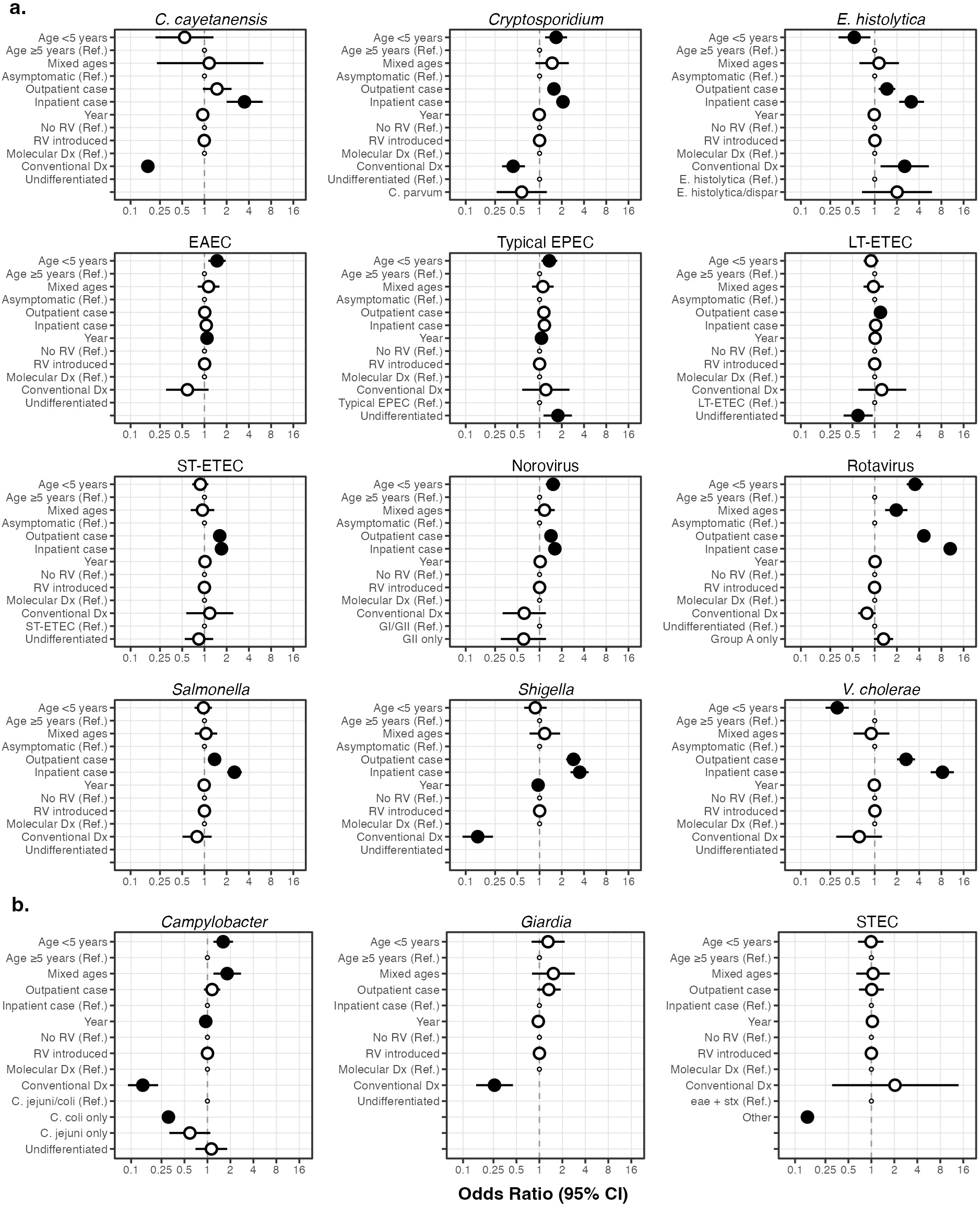
Dot-and-whisker plots of the coefficient estimates and their 95% uncertainty intervals for all variables included in the 15 models (14 pathogens with STEC separated by LT- and ST- variants), separated by: a). Odds ratio approach - those for which the estimated odds of detection were higher in symptomatic than in asymptomatic cases; b). “Modified detection as etiology” approach - those for which that assumption did not hold. (RV = rotavirus vaccine, Dx = diagnostic, Ref. = referent category).

Being in the younger age group (<5 years) was associated with a statistically significantly increased odds of rotavirus (aOR = 3.52 [2.71, 4.54]), *Cryptosporidium* (1.67 [1.19, 2.36]), *Campylobacter* (1.61 [1.20, 2.16]), norovirus (1.52 [1.21, 1.90]), EAEC (1.48 [1.13, 1.94]), and EPEC (1.36 [1.06, 1.74]), and decreased odds of *V. cholerae* (0.31 [0.22, 0.45]) relative to those aged ≥5 years (**Figure 2**). The term for the temporal trend in the odds of detection (year) was only statistically significant for four of the pathogens, indicating decreasing prevalence over time for *Campylobacter* (0.95 [0.91, 0.99]) and *Shigella* (0.96 [0.92, 0.99]) and increasing for EAEC (1.08 [1.02, 1.15]) and EPEC (1.06 [1.02, 1.11]). The aORs for rotavirus vaccine having been introduced nationally at the time of data collection (included because it was assumed, *a priori*, to reduce the probability of a stool sample being positive for rotavirus and increase the corresponding probability for other pathogens) was in fact not statistically significant for any pathogen including rotavirus. The use of conventional diagnostic methods as opposed to PCR conferred a statistically significantly reduced odds of detection of *Campylobacter* (0.14 [0.09, 0.23]), *Shigella* (0.15 [0.09, 0.23]), *C. cayetanensis* (0.17 [0.03, 1.18]), *Giardia* (0.26 [0.15, 0.45]), and *Cryptosporidium* (0.44 [0.31, 0.63]), and only for *E. histolytica* was a statistically significant increased odds observed (2.55 [1.20, 5.42]).

After applying the respective methods to convert the aORs to PAFs, the 14 selected pathogens accounted for a larger combined PAF in the younger than the older age group and in the more, rather than the less, severe diarrheal syndrome – 85.4% in inpatient and 60.9% in outpatient cases aged <5 years compared to 64.6% and 47.6% respectively in those aged ≥5 years (**Figure 3**). In the under 5 years age group, roughly equal proportions of outpatient cases were attributable to viral and non-*E. coli* bacterial etiologies (19.0% and 18.6% respectively). However, while rotavirus dominated the former taxon (a PAF of 14.18% compared to 4.5% for norovirus), in the latter, *Campylobacter* and *Shigella* contributed roughly equally to the overall attributable burden (8.9% and 8.7% respectively). Protozoa were the next highest contributing taxon in the <5-year-old outpatient stratum (a combined PAF of 13.0%), with *Giardia* accounting for around two-thirds of this burden (PAF = 8.7%), *Cryptosporidium* around one-third (3.8%), and *E. histolytica* and *C. cayetanensis* contributing only negligibly (0.3% and 0.2% respectively). Among the *E. coli* pathotypes, which had a combined PAF of 10.2% in the <5 years outpatient group, ETEC was the largest contributor (6.0%), followed by STEC (2.3%) and EPEC (1.5%) with EAEC accounting for a negligible PAF (0.5%). The main difference explaining the lower overall PAF in outpatient cases aged ≥5 years was the reduced PAF for rotavirus (4.4%), and, to a lesser extent, *Campylobacter* (6.4%), although this was slightly offset by increases in the PAF for *Shigella* (10.2%) and *V. cholerae* (2.5%) compared to in the younger age group. The increase in the overall pathogen-attributable PAFs in inpatient cases compared to outpatient cases was largely due to the greater share of rotavirus in this syndrome, particularly in <5-year-olds, the largest PAF for any pathogen in any of the four age and syndrome strata (30.6%). However, an increased PAF was predicted for *Cryptosporidium* in same age group (9.1%), for *V. cholerae* in those ≥5 years (10.7%), and for EAEC (2.5% and 1.9%) and *Salmonella* (3.1% and 2.7%) in both age groups.

**Figure 3:**
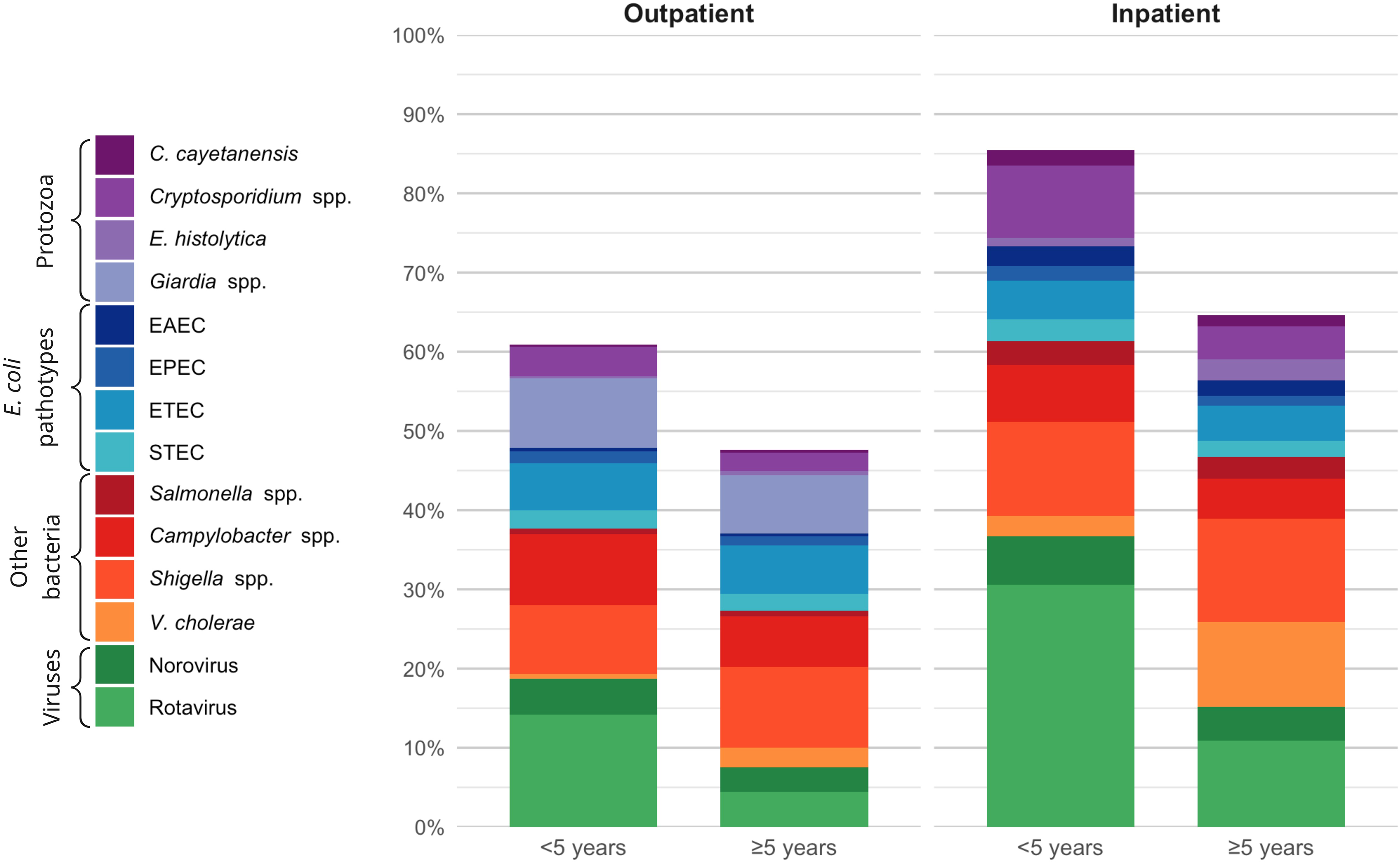
Etiology-specific population attributable fractions of 14 diarrhea-causing pathogens commonly transmissible via food in 2021 predicted by a meta-analytical modelling analysis arranged by taxonomical grouping.

**Table 2:** Numbers of studies, estimates and countries contributing to burden of illness incidence and mortality estimates in high income countries by pathogen.

| Pathogen | Incidence estimates (N=35 studies) |  |  | Mortality estimates (N=23 studies) |  |  |
| --- | --- | --- | --- | --- | --- | --- |
|  | Studies (N) | Estimates (N) | Countries (N) | Studies (N) | Estimates (N) | Countries (N) |
| *Campylobacter* spp. | 25 | 34 | 15 | 18 | 19 | 10 |
| *Cryptosporidium* spp. | 13 | 15 | 7 | 9 | 9 | 6 |
| *C. cayetanensis* | 3 | 3 | 2 | 3 | 3 | 2 |
| EAEC |  |  |  |  |  |  |
| *E. histolytica* | 1 | 1 | 1 | 0 | 0 | 0 |
| EPEC |  |  |  |  |  |  |
| ETEC | 4 | 4 | 3 | 3 | 3 | 2 |
| *Giardia* spp. | 12 | 12 | 6 |  |  |  |
| Norovirus | 19 | 20 | 9 | 16 | 17 | 8 |
| Rotavirus | 12 | 12 | 5 | 8 | 8 | 5 |
| *Salmonella* spp. | 29 | 36 | 14 | 20 | 21 | 9 |
| *Shigella* spp. | 13 | 18 | 11 | 11 | 11 | 8 |
| STEC | 18 | 22 | 10 | 13 | 14 | 7 |
| *V. cholerae* |  |  |  |  |  |  |

Shaded cells represent indicators that were not estimated due to the assumption that those pathogens are either not monitored, not endemic, or not a significant cause of mortality in HICs

### Approach 2 – Direct etiology-specific burden estimation for HICs

Approach 2 identified a much smaller initial set of publications (2,312), of which 98.4% (2,275) were excluded during screening, leaving 37 that were included in the analysis (**Figure 1a**). Of these, 35 studies contributed 42 sets of estimates on incidence for 1 or more of the 14 pathogens in 15 HICs and 23 studies contributed 24 sets of estimates for mortality for 1 or more of the 14 pathogens in 10 unique HICs. *Salmonella* had the largest number of estimates for incidence (39 from 14 countries), followed by *Campylobacter* (34 from 15 countries), STEC (22 from 10 countries), norovirus (20 from 9 countries), and *Shigella* (18 from 11 countries). ETEC (4 from 3 countries), *C. cayetanensis* (3 from 2 countries), and *E. histolytica* (1 from 1 country) had the fewest estimates. In general, that were fewer data points for mortality, but the trend was similar by pathogen in terms of the number of estimates available: *Salmonella* (21 from 9 countries), *Campylobacter* (19 from 10 countries), norovirus (17 from 8 countries), STEC (14 from 7 countries), and *Shigella* (11 from 8 countries). ETEC (4 from 3 countries), *C. cayetanensis* (3 from 2 countries), and *E. histolytica* (1 from 1 country) had the fewest estimates for incidence. 15 countries had one or more estimates for incidence or mortality: Australia, Barbados, Canada, Germany, Denmark, France, United Kingdom, Italy, Japan, Netherlands, New Zealand, Poland, Sweden, Switzerland, USA.

Bibliographic information for the publications identified by approaches 1 and 2 are provided as **Supplementary RIS files 1a and 1b**.

### Combined disease burden estimates

After applying the model-derived PAFs to the incidence and mortality envelopes for LMICs (approach 1) and adding them to the equivalent burden indicator values for HICs (approach 2), our estimates indicate that *Shigella* remains the pathogen responsible for the highest global diarrheal disease incidence, despite having almost halved by this measure between 2000 to 2021 (from 10,500 per 100,000 population in 2000 to 5,400 in 2021) (**Figure 4a**). As a cause of death, however the bacterium has declined the fastest of all the pathogens, such that in 2021, rotavirus had overtaken it as the leading etiology of diarrheal disease mortality. The next largest bacterial contributor to diarrheal disease morbidity (and the fastest declining of all the pathogens in terms of incidence) was *Campylobacter*. Declines in rotavirus disease incidence over the 21-year period were modest in comparison to the stark change in mortality due to this virus. In contrast, norovirus incidence was predicted to have increased slightly from 2,800 to 3,000 cases per 100,000 population despite this etiology declining as a cause of death from 1.4 to 0.8 per 100,000 population. *V. cholerae* also increased as a cause of morbidity, while exhibiting a mortality rate that decreased, though slowly, such that it was the third leading cause of diarrheal disease deaths by 2021 at 1 per 100,000. All four *E. coli* pathotypes increased in incidence over the period; two of them, EAEC and EPEC, showed increasing mortality rates in LMICs as well, with ETEC and STEC decreasing only modestly globally, such that, taken as a whole, pathogenic *E. coli* was the second and third leading etiology of diarrheal disease morbidity and mortality respectively in 2021.

**Figure 4:**
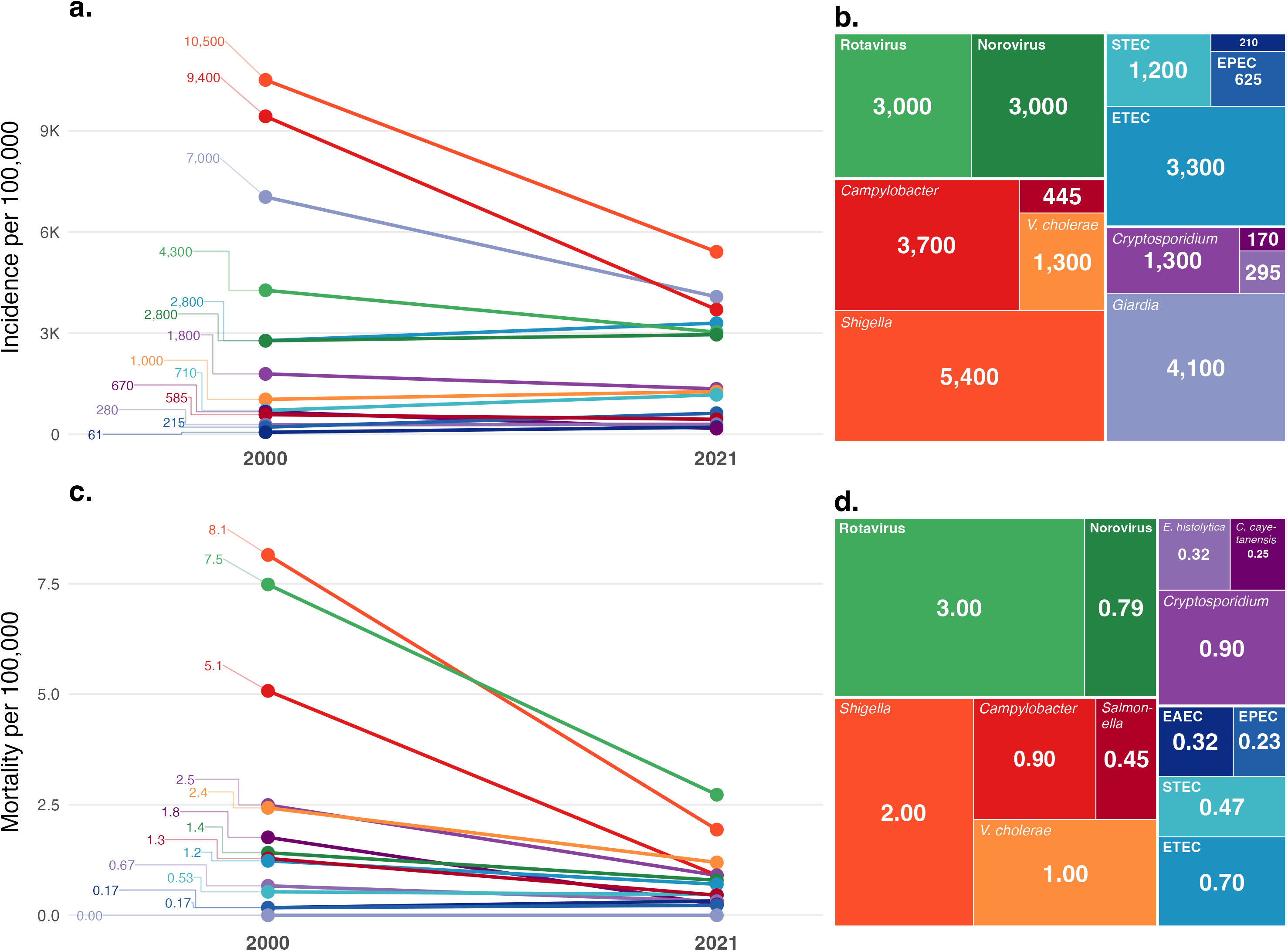
Changes in the predicted global diarrheal disease incidence (a.) and mortality (c.) rates per 100,000 population predicted by a meta-analytical modelling analysis for each of 14 pathogens between 2000 and 2021, with values for 2021 visualized as treemap plots (b. and d.).

Globally these 14 pathogens were responsible for 2.2 billion diarrheal disease cases and 880,000 deaths in 2021, with the largest number of cases occurring in the South-East Asian Region (SEAR), the largest number of deaths in the African Region (AFR), and Europe (EUR) being the region with the lowest burden by both measures (**Tables 3 and 4**). At national level, *Campylobacter* incidence was highest in India and Pakistan as well as numerous countries in Africa, notably those in the Sahel and Sudanian Savanna, as well as Angola and Madagascar (**Figure 5a**). Kenya was a striking outlier of low *Campylobacter* incidence. *Cryptosporidium* incidence did not exceed 6,000 per 100,000 outside of a handful of scattered countries in Africa, and was lowest in China, Mongolia, and several countries of Central Asia and the Balkans (**Figure 5b**). National level incidence of *C. cayetanensis* almost universally fell within the 10 – 500 per 100,000 range, with higher rates seen only in several Sub-Saharan African countries and lower in Mongolia, and several of the Balkan countries (**Figure 5c**). EAEC-attributable diarrhea is virtually absent from HICs and very low through most of the Americas and Asia, only exceeding 500 per 100,000 in Afghanistan and numerous Sub-Saharan African countries (**Figure 5d**). *E. histolytica* was not estimated for HICs and is also absent from HICs as well as Central Asia, China and Mongolia, Mexico, and several South American countries, and is highest in India, Pakistan, and some countries of the Sahel, Southern and East Africa, though still low compared to higher burden pathogens (**Figure 5e**). EPEC is present at low levels in HICs and exceeds 6,000 cases per 100,000 only in a handful of African countries (**Figure 5f**). ETEC- and *Giardia*- and *Shigella*-attributable diarrhea is endemic at high levels of incidence across Sub-Saharan Africa, India and Pakistan (**Figures 5g, 5h, and 5l**). Norovirus is the only pathogen with a morbidity burden in all HICs that is comparable to that in Sub-Saharan Africa, and greater than that found in South American, North African and Middle Eastern LMICs (**Figure 5i**). Rotavirus disease incidence exceeds 8,000 per 100,000 in Chad, South Sudan and Madagascar and has a higher incidence in HICs than in China and Mongolia. Salmonella has the highest incidence in the USA, Canada, Sub-Saharan Africa, India, Thailand and Papua New Guinea and lowest in Russia, China, Mongolia, Central Asia, and some countries of Eastern and Southeastern Europe (**Figure 5j**). STEC incidence is lowest in HICs outside of the Americas, and highest in Sub-Saharan Africa, India and Pakistan (**Figure 5m**), while *V. cholerae* is endemic only in Sub-Saharan Africa, South Asia, some Middle Eastern LMICs and the Philippines (**Figure 5n**).

**Figures 5a-n:**
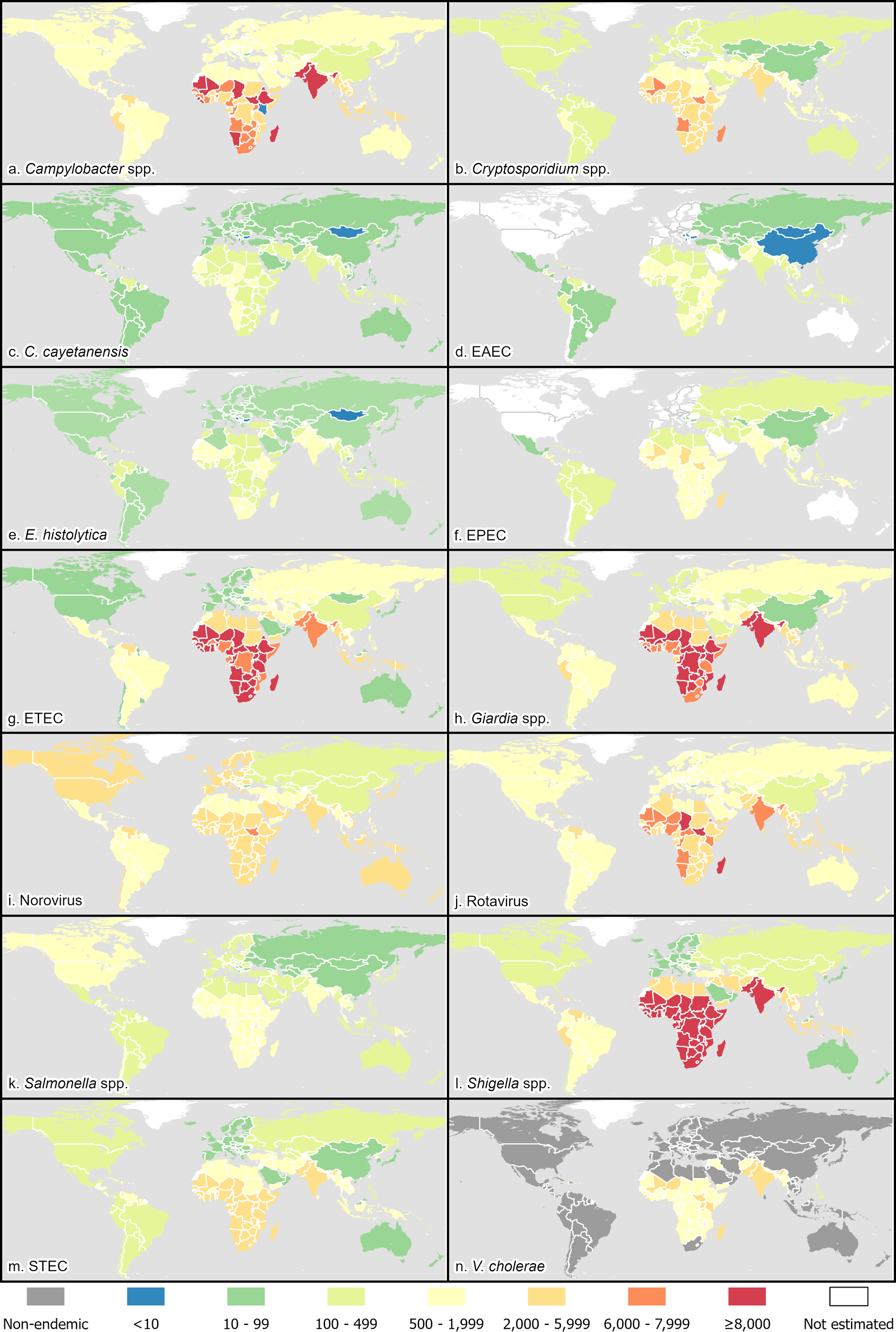
Incidence per 100,000 population of etiology-specific diarrheal disease in 2021 for 14 foodborne pathogens by country estimated by a meta-analytical modelling analysis.

**Table 3:** Annual etiology-specific diarrheal disease cases (thousands) for 14 foodborne pathogens in 2021 by WHO region and globally (with 95% uncertainty intervals) estimated from meta-analytic model, ranked by global total.

| Pathogen | African Region (AFR) | Region of the Americas (AMR) | Eastern Mediterranean Region (EMR) | European Region (EUR) | South-East Asian Region (SEAR) | Western Pacific Region (WPR) | Global |
| --- | --- | --- | --- | --- | --- | --- | --- |
| *Shigella* spp. | 137,800<br>(80,500, 220,500) | 11,400<br>(6,000, 19,200) | 46,200<br>(24,500, 83,100) | 2,800<br>(1,100, 5,700) | 220,300<br>(102,200, 407,000) | 7,800<br>(3,700, 14,000) | 426,400<br>(234,300, 711,400) |
| *Giardia* spp. | 115,000<br>(63,100, 188,200) | 8,600<br>(4,500, 14,500) | 36,800<br>(14,700, 69,700) | 4,600<br>(2,200, 8,100) | 147,800<br>(59,300, 315,000) | 8,400<br>(3,400, 16,400) | 321,200<br>(180,600, 581,700) |
| *Campylobacter* spp. | 81,300<br>(47,000, 126,400) | 11,400<br>(7,400, 16,500) | 33,000<br>(16,400, 57,400) | 7,200<br>(5,200, 9,900) | 146,800<br>(72,300, 250,300) | 11,700<br>(7,100, 18,300) | 291,400<br>(169,000, 458,600) |
| ETEC | 105,400<br>(62,100, 169,200) | 5,800<br>(3,000, 10,100) | 31,200<br>(16,200, 53,700) | 3,600<br>(1,700, 6,500) | 108,100<br>(55,900, 194,100) | 5,700<br>(2,900, 10,500) | 259,700<br>(151,700, 423,400) |
| Rotavirus | 68,900<br>(48,900, 97,700) | 12,300<br>(8,600, 17,500) | 21,900<br>(14,500, 31,900) | 8,800<br>(6,000, 13,000) | 110,100<br>(72,100, 177,000) | 16,600<br>(11,900, 22,500) | 238,600<br>(169,400, 339,200) |
| Norovirus | 45,300<br>(25,400, 71,200) | 29,400<br>(21,900, 37,600) | 20,100<br>(11,800, 30,300) | 33,500<br>(24,700, 43,000) | 82,900<br>(43,000, 137,900) | 21,700<br>(16,100, 28,500) | 232,800<br>(155,000, 332,000) |
| *Crypto-sporidium* spp. | 49,600<br>(25,900, 79,800) | 2,400<br>(1,200, 4,000) | 10,800<br>(5,100, 21,300) | 2,100<br>(1,000, 3,900) | 38,700<br>(17,900, 69,600) | 2,700<br>(1,200, 5,200) | 106,200<br>(57,400, 169,200) |
| *V. cholerae* | 16,600<br>(6,200, 37,700) | 24<br>(6, 61) | 10,100<br>(2,400, 27,800) | 0<br>(0, 0) | 72,800<br>(19,000, 166,800) | 245<br>(57, 630) | 99,800<br>(30,000, 227,800) |
| STEC | 34,100<br>(13,000, 76,300) | 2,800<br>(1,100, 6,100) | 10,300<br>(3,800, 23,400) | 2,100<br>(750, 5,000) | 41,000<br>(11,400, 117,600) | 2,000<br>(445, 5,200) | 92,400<br>(36,500, 204,900) |
| EPEC | 17,500<br>(510, 38,200) | 980<br>(28, 2,300) | 6,000<br>(165, 15,300) | 640<br>(13, 1,700) | 22,900<br>(550, 56,500) | 1,300<br>(25, 3,200) | 49,300<br>(1,300, 109,800) |
| *Salmonella* spp. | 9,700<br>(4,000, 18,500) | 3,600<br>(2,200, 5,400) | 2,600<br>(945, 5,400) | 2,000<br>(1,300, 3,000) | 14,400<br>(5,300, 31,500) | 2,700<br>(1,400, 4,600) | 35,000<br>(17,000, 63,600) |
| *E. histolytica* | 5,100<br>(1,200, 12,400) | 690<br>(175, 1,800) | 2,100<br>(385, 6,000) | 455<br>(105, 1,200) | 14,400<br>(2,300, 39,300) | 625<br>(150, 1,700) | 23,400<br>(4,800, 55,400) |
| EAEC | 5,600<br>(-61,500, 68,500) | 470<br>(-5,503, 6,000) | 2,000<br>(-20,942, 24,400) | 155<br>(-1,596, 2,000) | 7,800<br>(-85,907, 87,200) | 450<br>(-5,700, 6,100) | 16,600<br>(-174,679, 191,500) |
| *C. cayetanensis* | 7,200<br>(-561, 26,100) | 395<br>(2, 1,300) | 1,400<br>(-117, 5,300) | 245<br>(40, 860) | 3,700<br>(-293, 14,000) | 485<br>(-16, 2,000) | 13,400<br>(-1,059, 42,300) |
| **Total** | 699,100<br>(547,800, 870,000) | 90,300<br>(74,200, 108,000) | 234,500<br>(173,600, 316,900) | 68,300<br>(56,100, 81,200) | 1,031,800<br>(756,800, 1,376,900) | 82,300<br>(65,800, 101,100) | 2,206,300<br>(1,738, 200, 2,749,000) |

**Table 4:** Annual etiology-specific diarrheal disease deaths for 14 foodborne pathogens in 2021 by WHO region and globally (with 95% uncertainty intervals) estimated from meta-analytic model, ranked by global total.

| Pathogen | African Region (AFR) | Region of the Americas (AMR) | Eastern Mediterranean Region (EMR) | European Region (EUR) | South-East Asian Region (SEAR) | Western Pacific Region (WPR) | Global |
| --- | --- | --- | --- | --- | --- | --- | --- |
| **Rotavirus** | 109,800<br>(81,700, 143,200) | 3,200<br>(2,300, 4,300) | 19,600<br>(13,600, 26,600) | 785<br>(550, 1,200) | 75,300<br>(47,900, 109,900) | 6,000<br>(4,300, 7,800) | 214,700<br>(162,100, 275,300) |
| ***Shigella* spp.** | 62,700<br>(38,000, 94,600) | 2,100<br>(1,000, 3,400) | 13,800<br>(7,000, 24,400) | 240<br>(96, 470) | 72,500<br>(33,700, 127,100) | 1,200<br>(545, 2,300) | 152,500<br>(85,300, 242,300) |
| ***V. cholerae*** | 17,500<br>(7,700, 33,400) | 68<br>(18, 170) | 7,300<br>(2,200, 17,500) | 0<br>(0, 0) | 69,100<br>(21,800, 148,200) | 150<br>(35, 350) | 94,100<br>(34,200, 186,100) |
| ***Campylobacter* spp.** | 29,100<br>(15,300, 47,900) | 1,200<br>(670, 2,000) | 7,700<br>(3,500, 14,900) | 1,100<br>(565, 1,700) | 30,900<br>(13,400, 56,400) | 1,100<br>(540, 2,000) | 71,200<br>(37,200, 117,900) |
| ***Crypto-sporidium* spp.** | 44,700<br>(25,100, 70,600) | 525<br>(280, 935) | 5,900<br>(2,700, 11,600) | 180<br>(98, 300) | 19,100<br>(9,100, 35,100) | 610<br>(235, 1,300) | 71,000<br>(40,700, 111,600) |
| **Norovirus** | 24,700<br>(13,900, 39,700) | 1,800<br>(1,100, 2,800) | 5,800<br>(3,100, 9,700) | 2,000<br>(995, 3,300) | 26,400<br>(13,600, 43,700) | 1,600<br>(960, 2,600) | 62,300<br>(36,700, 95,400) |
| **ETEC** | 27,900<br>(13,900, 48,100) | 570<br>(230, 1,100) | 5,400<br>(1,900, 10,500) | 125<br>(41, 285) | 20,800<br>(7,700, 44,700) | 510<br>(165, 1,100) | 55,300<br>(27,500, 102,300) |
| **STEC** | 17,500<br>(7,100, 36,600) | 795<br>(345, 1,600) | 3,300<br>(1,100, 7,500) | 530<br>(240, 1,200) | 14,300<br>(4,100, 36,800) | 460<br>(135, 1,200) | 36,900<br>(15,400, 75,300) |
| ***Salmonella* spp.** | 15,500<br>(8,100, 28,600) | 1,100<br>(730, 1,600) | 2,400<br>(1,200, 4,300) | 900<br>(585, 1,400) | 14,800<br>(7,000, 29,400) | 1,000<br>(645, 1,700) | 35,800<br>(21,100, 60,400) |
| **EAEC** | 11,600<br>(-18,624, 47,300) | 400<br>(-650, 1,600) | 2,600<br>(-4,306, 10,900) | 58<br>(-107, 265) | 10,500<br>(-16,951, 44,200) | 360<br>(-603, 1,700) | 25,500<br>(-39,717, 104,200) |
| ***E. histolytica*** | 6,600<br>(2,300, 13,800) | 315<br>(87, 805) | 1,800<br>(500, 4,400) | 55<br>(13, 155) | 16,200<br>(4,200, 41,300) | 235<br>(50, 650) | 25,100<br>(7,300, 58,300) |
| ***C. cayetanensis*** | 12,900<br>(3,100, 35,900) | 235<br>(39, 755) | 1,300<br>(270, 3,700) | 38<br>(5, 120) | 4,700<br>(765, 16,900) | 220<br>(9, 830) | 19,300<br>(4,400, 58,300) |
| **EPEC** | 8,500<br>(-1,519, 22,700) | 175<br>(-24, 485) | 1,900<br>(-292, 4,900) | 46<br>(-7, 140) | 7,000<br>(-990, 19,500) | 215<br>(-28, 615) | 17,800<br>(-2,946, 49,500) |
| **Total** | 389,000<br>(321,700, 471,100) | 12,400<br>(10,100, 15,200) | 78,700<br>(58,300, 101,800) | 6,000<br>(4,700, 7,700) | 381,500<br>(279,500, 519,700) | 13,800<br>(10,900, 17,100) | 881,400<br>(716,900, 1,081,300) |

## Discussion

In this article we report updated estimates of pathogen-attributable diarrheal disease incidence and mortality at national, regional and global levels, that will serve as inputs for the WHO’s broader estimates of hazard-specific burden from foodborne diseases. These findings come at a pivotal time in the effort to control diarrheal disease, as the global health community takes stock of both the enormous progress made in reducing the most severe outcomes of this syndrome, as well as the persistent challenges that lie ahead in controlling these infections worldwide. It had long been assumed that low-cost, onsite WASH interventions, such as point-of-use water treatment and ventilated pit latrines (VIP), offered solutions that could substitute for more sweeping investments in municipal-level water, wastewater and drainage infrastructure of the kind that historically engendered society-wide child health improvements in HICs [34,35]. However, several recent highly powered trials [8–10] found mixed, negligible or no impacts of household WASH and nutrition interventions on diarrhea outcomes. Community- [36,37] and neighborhood-level [38] WASH interventions have shown similarly mixed results and are logistically and financially costly. It is possible that the historical focus on all-cause diarrhea outcomes masks pathogen-specific differences in the influence of intra- and extra-domiciliary environmental factors that derive from distinct ecological sensitivities of specific enteric pathogen species [39,40]. Furthermore, the effectiveness of decentralized household sanitation systems in preventing pathogen exposure may be limited in settings with poor drainage infrastructure that are easily overwhelmed by heavy rainfall events [41–43]. Meanwhile, the success of the rotavirus vaccine has prompted vaccine developers to target other high-burden enteropathogens, including other enteric viruses such as norovirus [44,45] as well as bacterial agents - *Shigella*, *Campylobacter*, and ETEC [46–48] – though progress has been slow. Other pathogen- and taxon-specific interventions, such as appropriate use of antibiotics to reduce case fatality [49], livestock management [50,51], and pre-harvest and production chain interventions to reduce foodborne bacteria transmission [52,53], are likely to be key strategies for sustaining progress, making etiology-specific burden estimates like these timely and important for prioritizing intervention targets.

The decade since the WHO’s FERG published the previous round of estimates has seen an increasing interest in and evidence about the profile of enteropathogens in diarrheal disease cases and how it compares to that of asymptomatic individuals [19]. This has in part been engendered by the emergence of multiplex molecular diagnostic platforms such as the Taqman Array Card [54], and BioFire FilmArray GI Panel [55], as well as the findings of several rigorously designed and well-resourced, multi-site studies in which these technologies have been deployed [7,56,57]. This is reflected in the large number of data sources identified as relevant by the broad-scope systematic review component of this study, with some 324 studies contributing data to the meta-analysis. In this iteration we employ several methodological advances compared with the 2015 one. Firstly, we consider an expanded panel of pathogens, including rotavirus, *C. cayetanensis* and EAEC for the first time. Secondly, all 14 pathogens were incorporated within the same systematic reviews, whereas previously they had been integrated from several independent syntheses of different combinations of pathogens. Thirdly, we developed and deployed an advanced extraction template capable of capturing the hierarchical nature of data reported in multiple strata, nested within sites and studies, as well as relevant covariates that were previously not considered, such as diagnostic method, pathogen strain, and vaccine introduction [19]. Fourthly, we used the now favored OR approach to calculating PAFs of pathogens for which the relevant assumptions were met, allowing us to adjust estimates for background prevalence of infection in individuals not exhibiting overt clinical symptoms.

We also diverge methodologically from several other recent meta-analytical studies that have addressed similar research questions. The GBD 2021 Diarrhoeal Diseases Collaborators (GBD) also sourced positivity rates from a systematic review for an overlapping group of 13 pathogens (not restricted to those capable of foodborne transmission) to derive etiology-specific PAFs from a meta-analytical model, however, they only extracted data for symptomatic subjects, and adjusted these for ORs for disease that were themselves calculated from just two studies, MAL-ED and GEMS, conducted in young children in 15 LMIC sites. Black et al. used a similar approach to a slightly different group of 12 pathogens diagnosed in hospitalized children, also using ORs from the same two studies, but that took into account pathogen quantities derived from the PCR quantification cycle values [58]. By contrast, we derive ORs by comparing symptomatic detection rates with asymptomatic ones taken from a larger, collective set of diverse and more geographically representative studies, with random effects to specify clustering of these strata within the same study and site, thus overcoming any potential biases associated with using adjustments based on just 15 locations (the 8 MAL-ED and 7 GEMS sites) and in the <5 years age group. Other comparable studies to ours have adopted the DE approach to calculating PAFs and restricted to particular regions [59], pathogens [60,61] or taxa [62,63].

Perhaps as a consequence of these modifications, our findings differ in important ways from those of previous estimation attempts. We found the leading causes of diarrhea morbidity to be bacterial – *Shigella*, *Campylobacter* and ETEC – and protozoal – *Giardia* – agents, contradicting a large body of evidence that places viruses, notably rotavirus, at the top of the ranking [7,64]. The first FERG analysis did not include rotavirus, but estimated norovirus to be the largest contributor to diarrhea incidence in 2010 by some margin (responsible for almost 3 times the number of illness of the next pathogen, ETEC) [18]. Even when taking the estimates for 2010 from this study for comparison, norovirus comes in at a distant 7^th^ place with 36 million annual cases estimated for that year compared to the previously estimated 685 million, a difference that is likely explained by the replacement of the DE with the OR approach for this pathogen [18]. Norovirus shedding is prolonged following infection, and virions are detectable in stool for several weeks after symptoms have resolved [65]. This residual shedding coupled with the high force of infection results in a substantial background prevalence of norovirus in asymptomatic individuals. In this study, a pooled 12.2% of asymptomatic subjects in LMICs were norovirus-positive (**Table 1**) and elsewhere this rate has been estimated at 7% globally, as high as 15% in Africa, and 16% when considering only PCR diagnostic methods [66] (the predominant modality represented in our dataset). It is possible that this widespread presence of norovirus in stool samples from individuals in whom it is no longer eliciting clinical symptoms biased our aORs towards the null, leading to an underestimation of that pathogen’s true PAF.

GBD estimated rotavirus to be the largest contributor to diarrheal disease burden (though in terms of DALYs, not incidence), followed by *Shigella*, adenovirus (not considered foodborne and so excluded from this study) and *Cryptosporidium* (7^th^ by cases, 5^th^ by deaths in our results) [64]. Our updated estimates support a much higher morbidity burden for *Shigella* than previously - 506 million annual cases in 2010 globally compared with 190 million [15] – with an incidence rate for 2021 that amounts to more than one in every 20 people suffering an episode of shigellosis. GBD put *Shigella* as the second highest by DALYs in all ages, however, in the 7-site GEMS study, and the VIDA follow-up study conducted in the 3 of the African GEMS sites following introduction of the rotavirus vaccine, *Shigella* was found to have the highest PAF among children <5 years with moderate-to-severe diarrhea (a PAF of 19.8% compared to 12.6% for rotavirus in VIDA) [56,67,68]. In fact it is the larger PAF for *Shigella* in outpatients aged ≥5 years (10.2%) that seems to account for the higher overall shigellosis incidence and mortality in our estimates. Regarding etiology-specific death attribution, our estimates align with GBD [64], Black et al. [58] and the Global Pediatric Diarrhea Surveillance (GPDS) [7] study in ranking rotavirus first, and the latter two of those (which only estimated them for <5s) in putting *Shigella* second. Our ranking of *V. cholerae* as third largest mortality etiology is likely due to that bacterium’s relatively large PAF in inpatients aged ≥5 years, cholera being a disease for which high rates of severe disease and case-fatality are observed in adults but less so in children [69,70].

In attributing a large proportion of diarrheal disease morbidity to giardiasis, our findings are at odds with numerous previously reported studies. While we excluded *Giardia* from mortality estimations, GBD also excluded the pathogen from incidence calculations [64], and many high-profile observational etiology attribution studies either do not test samples for or do not report *Giardia* positivity [7,56]. While it is beyond doubt that *Giardia* is a direct cause of diarrheal illness [71], particularly initial and persistent infections occurring in young, immunologically naïve infants in LMICs [72], the parasite is also found at high levels of prevalence in later childhood usually in the absence of concurrent symptoms (though intestinal permeability and linear growth faltering are among its longer term sequalae) [72]. This is a consequence of the protozoa’s tendency to elicit rapid acquisition of long-lasting immunity and prolonged infections lasting months [73,74], as well as the generally high force of infection in low-resource settings with inadequate access to WASH infrastructure. Our finding of an inverse association between diarrheal symptom severity and the probability of *Giardia* detection is congruent with a phenomenon that is well-documented in observational [54,75] and meta-analytical studies [76,77]. The strength and consistency of this association (pooled ORs of 0.4 [76] to 0.6 [77]) has led some commentators to speculate that it is due to a genuine protective effect whereby the presence of chronic *Giardia* infection in the gut attenuates clinical symptoms or modulates the immune response to concurrent infections with other pathogens [78,79]. We consider a more likely explanation to be that in cases of watery diarrhea (or the steatorrhea characteristic of giardiasis [80]) the density of parasites in the stool is diminished due to purging and more likely to fall below the diagnostic threshold [81]. The implication is that neither the OR nor the DE method of PAF calculation are adequate for estimation of giardiasis burden, with the assumptions of the former not being met by the epidemiology of the pathogen and the latter vastly overestimating the symptomatic proportion. With our mDE approach we took the average of the two extremes of the range of PAFs that were consistent with the data, which essentially amounted to halving the PAF as estimated by conventional DE.

Perhaps a more novel finding of our study is that *Campylobacter* followed a similar pattern to *Giardia*, with pooled prevalence in both outpatient and inpatient diarrhea cases much lower than in asymptomatic subjects leading to modeled ORs of less than 1. Although we adjusted for diagnostic method in the multivariable model (an adjustment that led to plausible model outputs for other pathogens), it is possible that this was due to residual confounding by this variable in the case of *Campylobacter*. It is notable that a single large study from China, which used culture on samples from >90,000 diarrheal cases, made up a large proportion (almost one third) of the outpatient cases and only reported a *Campylobacter* prevalence of <1%, a data point that could have skewed the overall result [82]. However, even when we excluded results diagnosed by culture in a sensitivity analysis and only considered PCR results, the differential reversed only in outpatients, with inpatient cases still showing around half the *Campylobacter* positivity of asymptomatic cases (**Supplementary table S1**). To assess the potential for confounding by heterogeneity in study design, we made the same comparison using PCR results only from the GEMS study, and found that while the effect attenuated, prevalence in GEMS cases that were admitted as inpatients was still 6.6 percentage points lower than in GEMS controls, while prevalence in outpatient cases was 3.8% higher [56]. This divergence was not noted in the original GEMS publications in which the authors pooled all MSD cases together regardless of severity [56,83] and found positive PAFs for *Campylobacter* (though with some negative ones in specific sites [56]), however, by analyzing the study’s microdata, we were able to make this distinction in our analysis and use GEMS data for all three syndrome strata in our analysis. It is unlikely that this phenomenon is due to antibiotic use, since in most studies protocols specify taking samples before administration of antibiotics [47,84]. More likely it is due to the accumulation of acquired immunity protecting against symptomatic campylobacteriosis but not infection, thus inflating the denominator in the OR calculation with subjects who are not at risk of disease and biasing it towards – or in this case across - the null [85]. Furthermore, molecular diagnostics for *Campylobacter* remain sub-optimal, failing to reliably detect all *Campylobacter jejuni*/*coli* due to primer selection based on a set of genomes isolated from HIC contexts. While PCR has improved sensitivity relative to culture, the use of probes that are broadly reactive across non-pathogenic species is likely to lead to a misclassification of attribution. STEC too had an OR for disease of <1 due to a larger prevalence of detection in asymptomatic than in symptomatic individuals, though given the low overall transmission level and the relatively few studies that tested for this pathogen, this could be due to random error.

Our study has limitations. Because the scope was limited to pathogens capable of foodborne transmission, we did not estimate the burden of important diarrhea-causing enteropathogens largely transmitted by other routes such as adenovirus, astrovirus, sapovirus, and *Clostridium difficile*, thus limiting comparisons with other similar efforts. For consistency with the earlier iteration of this study, we used different approaches for HICs and LMICs, however, the current availability of high-quality evidence from both geographies makes this distinction much less relevant than it was a decade ago. Systematic review approaches to diarrheal disease etiology attribution such as this cannot account for coinfections with multiple pathogens, which occur extremely frequently [86] and may cause misattribution of attribution or have multiplicative effects on case severity such that no single pathogen is the sole cause of some episodes. A tacit assumption of approach 1 is that studies carried out in single locations are representative of the entire country, whereas in reality this is seldom the case. We pooled community-detected diarrheal episodes (such as those in MAL-ED) with the far more numerous outpatients treated cases (such as GEMS cases) thus skewing our incidence estimates towards more severe cases. Furthermore, many studies did not report outpatient and inpatient cases separately. In such cases, we classified them as outpatient in order not to down-bias mortality estimates, but as a consequence may have inflated PAFs for outpatient cases. The PAF approach itself has limitations that are exemplified by this analysis, such as that pathogens that elicit prolonged residual shedding following resolution of symptoms (e.g., norovirus, *Campylobacter*, *Giardia*) will have large asymptomatic proportions that can artificially attenuate their PAF estimate.

Our classification of age into two broad groups may mask important shifts in the age-specific pathogen profile of diarrhea, particularly over the first few years of life [57]. Norovirus incidence is highest in the 6 – 11 month age range [87], while *Shigella* comes to dominate in later childhood [88] so it is possible that the apparent down-ranking of the former relative to the latter in our estimates is the result of this overly coarse age classification. However, most studies do not report results at the level of disaggregation which would allow for a more highly granular age specific attribution. This would be a valuable future improvement in these estimates in the future and should inform best practices for reporting in observational studies. More generally, there is a limited body of evidence for older age groups. Finally, it should also be considered that certain pathogens, such as *Cryptosporidium,* have heavy burdens in specific high-risk groups (e.g. malnourished, immuno-compromised) and aggregate global estimates based on typical risk levels may underestimate the burden in these more susceptible groups.

In conclusion, we present updated pathogen-specific incidence and mortality estimates for global etiology of acute diarrheal diseases compared over a 22-year period. These estimates are supported by a substantial increase in the available sites contributing data which significantly strengthens both global and regional estimates for policy makers.

Additionally, the more comprehensive set of pathogens sought often using more sensitive molecular diagnostic methods increases the reliability of these estimates. There remains a need for refined statistical methods to accurately quantify PAFs for pathogens such as norovirus and campylobacteriosis, that are recognized as clinically important pathogens globally, but violate the assumptions of the OR method due to their tendency for prolonged asymptomatic carriage and infection following resolution of symptoms relative to other pathogens. Despite their limitations, these improved etiology-specific estimates can be used to generate global burden of foodborne disease estimates and can inform the value proposition of vaccine development and food processing and packaging in attempts to limit the transmission of those agents with the greatest disease burden.

## Acknowledgements

The following people assisted with obtaining and translating articles that were in languages not spoken by the primary authors (English, Spanish and Japanese): Ebru Aydin, M. Kirami Ölgen, and Gültekin Unal (Turkish); Ruthly Francois (French); Rafal Gierczyński (Polish); Yunchang Guo (Chinese); Mirjam Kretzschmar (German); Lapo Mughini Gras (Italian); Ali Rostami (Persian); Michaela Špačková (Czech); Soohwan Suh (Korean); Paul Torgerson (Russian); Theodoros Varzakas (Greek); Pablo Yori (Catalan). The following people allowed the investigators access to their study datasets to calculate stratified aggregate aetiology proportions: Ayola Akim Adegnika; Amanda Debes; Poul-Erik Lund Kofoed; Ralf Krumkamp; Nina Langeland; Aldo Lima; Prince Manouana; Sabrina Moyo; Kazuhisa Okada; Nicola Page; James Platts-Mills; Firdausi Qadri. Bochen Cao at WHO provided diarrhea mortality data that were used as envelopes. The following staff at the Institute for Health Metrics and Evaluation facilitated access to draw-level data estimates that were used as the diarrhea incidence envelopes: Hmwe Kyu, Amanda Novotney, Kelly Bienhoff and Rafael Lozano. The following people provided expert consultation on the methods and findings: Robert Black; Paul Hunter; Benjamin Lopman; Elizabeth Rogawski-McQuade. Staff of the University of Virginia’s Claude Moore Health Sciences Library provided consultation on systematic review methods (Andrea Denton and Dan Wilson) as well as facilitating countless inter-library loans of obscure articles. Malena Nong provided data visualization support early in the project. Sierra Meyer assisted with updating the scoping review.

## Ethical standards

All human subject information used was aggregated secondary data. For studies for which individual level microdata was used, these were either already shared with the authors under data use agreements and used with specific permission for this purpose, or otherwise publicly available. We followed the PRISMA, GATHER and STROBOD guidelines (**supplementary file S2**).

## Data availability

A database of our full estimates stratified by year, age group, sex, country, and burden indicator, as well as the PAFs in figure 3, is available as **supplementary file S3**. The extracted data to which models were fitted is available upon reasonable request to the communicating author.

## Supporting Information

**Supplementary files S1a and S1b: RIS file of bibliographical information for publications identified as eligible for inclusion by approaches 1 and 2.**

**Supplementary file S2: Supplementary methods, results, and reporting checklists.**

**Supplementary file S3: Excel file of full estimates stratified by year, age group, sex, country, and burden indicator**

