## Supplementary file S2 for "Updated Estimates of the Global, Regional and National Burden, and Etiology of Diarrheal Diseases Transmissible via Food: A Systematic Review and Meta-Analytical Modelling Study for the World Health Organization"

### Search strategies for Approach 1 - Indirect etiology attribution for LMICs:

##### *Medline - PubMed*

((((((("diarrhea*"[tiab] OR "diarrhoea*"[tiab] OR "gastroenteritis"[tiab] OR "Dysentery"[tiab] OR ("feces/microbiology"[mh] OR "feces/parasitology"[mh] OR "feces/virology"[mh]) OR "Dysentery"[mh]) AND ("campylobacter*"[tiab] OR "campylobacter"[mh] OR ("cryptosporidi*"[tiab] OR "cryptosporidium"[mh]) OR ("Cyclospora"[tiab] OR "Cyclosporiasis"[tiab] OR "Cyclospora"[mh]) OR ("enteroaggregative e coli"[tiab] OR "Enteroaggregative Escherichia coli"[tiab] OR "EAEC"[tiab] OR "EAggEC"[tiab]) OR ("entamoeba"[tiab] OR "entameba"[tiab] OR "entamebiasis"[tiab] OR "entamoebiasis"[tiab] OR "amebiasis"[tiab] OR "amoebiasis"[tiab] OR "amoebic dysentery"[tiab] OR "amebic dysentery"[tiab] OR "entamoeba histolytica"[mh]) OR ("enteropathogenic e coli"[tiab] OR "Enteropathogenic Escherichia coli"[tiab] OR "EPEC"[tiab] OR "Enteropathogenic Escherichia coli"[mh]) OR ("enterotoxigenic e coli"[tiab] OR "enterotoxigenic escherichia coli"[tiab] OR "ETEC"[tiab] OR "enterotoxigenic escherichia coli"[mh]) OR ("giardia*"[tiab] OR "giardia"[mh]) OR ("norovirus"[tiab] OR "Norwalk virus"[tiab] OR "Norwalk agent"[tiab] OR "norovirus"[mh]) OR ("rotavirus"[tiab] OR "rotavirus"[mh]) OR ("Salmonella enterica"[tiab] OR "paratyphoid fever"[tiab] OR "typhoid fever"[tiab] OR "Salmonella enterica"[mh]) OR ("shigella"[tiab] OR "shigellosis"[tiab] OR "bacillary dysentery"[tiab] OR "shigella"[mh]) OR ("shiga toxin producing e coli"[tiab] OR "Shiga toxin-producing Escherichia coli"[tiab] OR "shiga toxigenic e coli"[tiab] OR "shiga toxigenic escherichia coli"[tiab] OR "STEC"[tiab] OR "shiga toxigenic escherichia coli"[mh]) OR ("Vibrio cholerae"[tiab] OR "cholera"[tiab] OR "Vibrio cholerae"[mh]))) OR ("enteropathogens"[ti] OR "enteropathogen"[ti] OR "enteric pathogens"[ti] OR "enteric pathogen"[ti] OR "enteric infections"[ti] OR "enteroinfection*"[ti])) AND 1990/01/01:2023/06/30[dp]) NOT ("animals"[mh] NOT "humans"[mh])) NOT ("pubmed books"[Filter] OR "meta analysis"[pt] OR "review"[pt] OR "systematic review"[Filter])) AND ("all"[Filter] NOT "preprint"[pt])

##### *Web of Science*

(((TS=("diarrhea*" OR "diarrhoea*" OR "gastroenteritis" OR "dysentery") AND TS=("campylobacter*" OR "cryptosporidi*" OR "cyclospora" OR "cyclosporiasis" OR "Enteroaggregative E. coli" OR "Enteroaggregative Escherichia coli" OR "EAEC" OR "EAggEC" OR "entamoeba" OR "entameba" OR "entamebiasis" OR "entamoebiasis" OR "amebiasis" OR "amoebiasis" OR "amoebic dysentery" OR "amebic dysentery" OR "entamoeba histolytica" OR "Enteropathogenic E. coli" OR "Enteropathogenic Escherichia coli" OR "EPEC" OR "Enteropathogenic Escherichia coli" OR "enterotoxigenic E. coli" OR "enterotoxigenic Escherichia coli" OR "ETEC" OR "Enterotoxigenic Escherichia coli" OR "giardia*" OR "norovirus" OR "Norwalk virus" OR "Norwalk agent" OR "rotavirus" OR "Salmonella enterica" OR "paratyphoid fever" OR "typhoid fever" OR "salmonella enterica" OR "shigella" OR "shigellosis" OR "bacillary dysentery" OR "Shiga toxin-producing E. coli" OR "Shiga toxin-producing Escherichia coli" OR "Shiga Toxigenic E. coli" OR "Shiga Toxigenic Escherichia coli" OR "STEC" OR "Vibrio cholerae" OR "cholera")) AND DOP=(1990-01-01/2023-06-30)) AND WC=((Gastroenterology & Hepatology) OR (Infectious Diseases) OR (Microbiology) OR (Parasitology) OR (Pediatrics) OR (Tropical Medicine) OR (Virology))) NOT DT=(Book OR Bibliography OR Biographical-Item OR Book Chapter OR Book

Review OR Chronology OR Correction OR Dance Performance Review OR Data Paper OR Database Review OR Discussion OR Editorial Material OR Excerpt OR Expression of Concern OR Fiction, Creative Prose OR Film Review OR Hardware Review OR Item About an Individual OR Item Withdrawal OR Music Performance Review OR Music Score OR Music Score Review OR News Item OR Note OR Poetry OR Review OR Script OR Software Review OR Theater Review OR TV Review, Radio Review OR TV Review, Radio Review Video)

***Embase***

[1990-2023]/py NOT [01/07/2023]/sd AND (diarrh*ea*:ab,ti OR gastroenteritis:ab,ti OR dysentery:ab,ti) AND (enteropathogens:ti OR enteropathogen:ti OR 'enteric pathogens':ti OR 'enteric pathogen':ti OR 'enteric infections':ti OR enteroinfection*:ti OR campylobacter*:ab,ti OR 'campylobacter'/exp OR cryptosporidi*:ab,ti OR 'cryptosporidium'/exp OR cyclospora:ab,ti OR cyclosporiasis:ab,ti OR 'cyclospora'/exp OR 'enteroaggregative e* coli':ab,ti OR eaec:ab,ti OR eaggec:ab,ti OR entam*eba:ab,ti OR entam*ebiasis:ab,ti OR am*ebiasis:ab,ti OR 'am*ebic dysentery':ab,ti OR 'entamoeba histolytica'/exp OR 'enteropathogenic e* coli':ab,ti OR epec:ab,ti OR 'enteropathogenic escherichia coli'/exp OR 'enterotoxigenic e* coli':ab,ti OR etec:ab,ti OR 'enterotoxigenic escherichia coli'/exp OR giardia*:ab,ti OR 'giardia'/exp OR norovirus:ab,ti OR 'norwalk virus':ab,ti OR 'norwalk agent':ab,ti OR 'norovirus'/exp OR rotavirus:ab,ti OR rotavírus OR 'salmonella enterica':ab,ti OR 'paratyphoid fever':ab,ti OR 'typhoid fever':ab,ti OR 'salmonella enterica'/exp OR shigella:ab,ti OR shigellosis:ab,ti OR 'bacillary dysentery':ab,ti OR 'shigella'/exp OR 'shiga toxin-producing e* coli':ab,ti OR 'shiga toxigenic e* coli':ab,ti OR stec:ab,ti OR 'shiga-toxigenic escherichia coli'/exp OR 'vibrio cholerae':ab,ti OR cholera:ab,ti OR 'vibrio cholerae'/exp) NOT ('animals'/exp NOT 'humans'/exp) NOT ('review':it OR 'systematic review':it OR 'meta-analysis':it OR 'book':it OR 'editorial':it OR 'preprint':it OR 'news':it OR 'patent':it)

#### Detailed modeling methods

Overall, the methodology followed the general methodology developed for the WHO estimates of the global burden of foodborne disease by the FERG Computational Task Force (ref CTF paper).

##### *Hierarchical meta-regression model*

We adopted a hierarchical meta-regression model for pooling, smoothing and imputing data from the systematic reviews, and generating internally consistent estimates for all countries from 2000 to 2021.

##### *Approach 1 - Indirect etiology attribution for LMICs:*

For the etiology prevalence data, a more elaborate model was applied that allow for more detailed geographical clustering and for the inclusion of additional study-level covariates. The model was applied to the logit-transformed prevalence data and corresponding standard errors, obtained using the escalc function from the metafor package. The model was specified as follows:

$$\hat{\theta}_{ijklm} = \theta + \beta_y . year_{ijklm} + \boldsymbol{\beta X}_{ijklm} + \zeta_{(6)m} + \zeta_{(5)lm} + \zeta_{(4)klm} + \zeta_{(3)jklm} + \zeta_{(2)ijklm} + \epsilon_{ijklm}$$

Where:

- $\hat{\theta}_{ijklm}$ is an estimate of the true effect size $\theta_{ijklm}$ for data point $i$ nested in study $j$, nested in country $k$, nested in subregion $l$, nested in region $m$.
- $\theta$ is the global intercept
- $\beta_y$ is the regression coefficient for the global effect of year
- $\boldsymbol{\beta}$ is the regression coefficients for possible global covariates $\boldsymbol{X}$, including syndrome type (asymptomatic, outpatient, inpatient), age group (below 5, above 5, mixed age groups), diagnostic method (molecular, other), rotavirus vaccination coverage, and, as relevant, strain
- $\zeta_{(6)m}$ is the random effect for region $m$
- $\zeta_{(5)lm}$ is the random effect for subregion $l$, nested in region $m$
- $\zeta_{(4)klm}$ is the random effect for country $k$, nested in subregion $l$, nested in region $m$
- $\zeta_{(3)jklm}$ is the random effect for study $j$ nested in country $k$, nested in subregion $l$, nested in region $m$

- $\zeta_{(2)ijklm}$ is the random effect for data point $i$ nested in study $j$, nested in country $k$, nested in subregion $l$, nested in region $m$
- $\epsilon_{ijklm}$ is the residual error term, pre-defined based on the study sample size.

Based on the fitted models, etiological fractions were estimated for incident cases and deaths, for two broad age groups (<5; ≥5) and both sexes, for the year 2000 through 2021, per country.

For a majority of pathogens, etiological fractions were calculated as attributable fractions, comparing the prevalence in asymptomatic vs symptomatic individuals:

$$EF = P_s * (1 - 1/OR)$$

$$OR = \frac{P_s/(1 - P_s)}{P_a/(1 - P_a)}$$

For Campylobacter, Giardia, and STEC, the etiological fraction was estimated as the average between the prevalence in symptomatic individuals and zero.

##### *Approach 2 – Direct etiology-specific burden estimation for HICs:*

For the multiplier data from the national studies, a simplified version of the default model was implemented, given the sparsity of the data. Four models were applied to the log-transformed incidence and mortality data and corresponding standard errors, obtained using the escalc function from the metafor package. The full model contained regional clustering and a global time trend:

$$\hat{\theta}_{ij} = \theta + \beta_y.year_{ij} + \zeta_{(3)j} + \zeta_{(2)ij} + \epsilon_{ijk}$$

where $\hat{\theta}_{ijkl}$ is an estimate of the true log-transformed incidence rate $\theta_{ij}$ for study $i$ nested in region $j$. $\theta$ is the global intercept, and $\beta_y$ the regression coefficient for the global effect of year. The residual error term $\epsilon_{ijk}$ was fixed based on the sample size of the data point.

The simpler models omitted the regional random effect, the year fixed effect, or both. For each hazard-parameter, the best model was selected based on an algorithm that included the number of data points (no clustering if less than 10 datapoints), model convergence, and model fit.

##### *Imputation process:*

After fitting the hierarchical meta-regression model to the available data, posterior predictive distributions were used to impute values for countries with missing data. The imputation process followed a hierarchical structure:

1. If data were available for other countries in the same subregion, the subregion-year-specific estimate was used.
2. If the subregion lacked data but the broader region had data, the region-year-specific estimate was used.

3. If no data were available for the region, the global-year-specific estimate was applied.

For *Vibrio cholerae*, countries deemed free from exposure were excluded from the imputation model and assigned a zero etiological fraction.

##### *Bayesian implementation and convergence assessment:*

The models were implemented in a Bayesian framework to properly account for uncertainty arising from the estimation process. Specifically, we used the `brms` package in R, which interfaces with Stan. We used a Student-T(3) prior for the intercept, flat priors for the regression coefficients, and half-Normal(0,1) priors for the standard deviations of the random effects. We fitted five chains, each consisting of 20,000 iterations, of which the first 10,000 were discarded as warmup. This resulted in 50,000 retained iterations for each parameter. Parameter uncertainty was propagated via 10,000 draws of the posterior distributions. These 10,000 estimates were then summarized by their mean and a 95% uncertainty interval defined as the 2.5th and 97.5th percentile of the distribution of estimates.

Convergence was assessed by visually examining trace plots and be ensuring Rhat statistics to be close to one. We also assessed divergent transitions, and increased maximum tree depth in case of need.

### Supplementary results:

**Table S1:**

| | | **All** results | | | **PCR only** | | | GEMS-only | | |
|---|---|---|---|---|---|---|---|---|---|---|
| | | **Samples** | **Positives** | **Prevalence** | **Samples** | **Positives** | **Prevalence** | **Samples** | **Positives** | **Prevalence** |
| **Asymptomatic** | <12 months | 25,860 | 5,480 | 21.2% | 20,599 | 5,012 | 24.3% | 2,034 | 842 | 41.4% |
| | 12 - 23 months | 24,247 | 7,256 | 29.9% | 20,609 | 6,881 | 33.4% | 1,947 | 987 | 50.7% |
| | 24 - 59 months | 6,701 | 1,353 | 20.2% | 3,356 | 1,090 | 32.5% | 1,660 | 663 | 39.9% |
| | < 5 yrs | 73,093 | 15,146 | 20.7% | 45,854 | 13,157 | 28.7% | 5,641 | 2,492 | 44.2% |
| | >=5 yrs | 819 | 3 | 0.4% | 239 | 3 | 1.3% | - | - | - |
| | Total | 81,212 | 15,419 | 19.0% | 46,192 | 13,160 | 28.5% | - | - | - |
| **Outpatient** | <12 months | 38,418 | 3,043 | 7.9% | 7,838 | 2,075 | 26.5% | 1,564 | 712 | 45.5% |
| | 12 - 23 months | 8,835 | 2,489 | 28.2% | 5,428 | 2,142 | 39.5% | 1,537 | 842 | 54.8% |
| | 24 - 59 months | 5,427 | 1,011 | 18.6% | 2,007 | 736 | 36.7% | 1,341 | 579 | 43.2% |
| | < 5 yrs | 125,326 | 10,813 | 8.6% | 16,508 | 5,391 | 32.7% | 4,442 | 2,133 | 48.0% |
| | >=5 yrs | 20,463 | 528 | 2.6% | 653 | 70 | 10.7% | - | - | - |
| | Total | 289,086 | 16,872 | 5.8% | 23,458 | 5,929 | 25.3% | - | - | - |
| **Inpatient** | <12 months | 8,323 | 1,168 | 14.0% | 6,589 | 1,004 | 15.2% | 496 | 207 | 41.7% |
| | 12 - 23 months | 4,324 | 732 | 16.9% | 2,848 | 561 | 19.7% | 424 | 165 | 38.9% |
| | 24 - 59 months | 2,026 | 284 | 14.0% | 1,113 | 187 | 16.8% | 322 | 95 | 29.5% |
| | < 5 yrs | 22,199 | 2,607 | 11.7% | 11,960 | 1,924 | 16.1% | 1,242 | 467 | 37.6% |
| | >=5 yrs | 3,426 | 223 | 6.5% | 1,732 | 111 | 6.4% | - | - | - |
| | Total | 31,315 | 3,168 | 10.1% | 13,954 | 2,043 | 14.6% | - | - | - |
| **All diarrhea** | <12 months | 46,741 | 4,211 | 9.0% | 14,427 | 3,079 | 21.3% | 2,060 | 919 | 44.6% |
| | 12 - 23 months | 13,159 | 3,221 | 24.5% | 8,276 | 2,703 | 32.7% | 1,961 | 1,007 | 51.4% |
| | 24 - 59 months | 7,453 | 1,295 | 17.4% | 3,120 | 923 | 29.6% | 1,663 | 674 | 40.5% |
| | < 5 yrs | 147,525 | 13,420 | 9.1% | 28,468 | 7,315 | 25.7% | 5,684 | 2,600 | 45.7% |
| | >=5 yrs | 23,889 | 751 | 3.1% | 2,385 | 181 | 7.6% | - | - | |
| | Total | 320,401 | 20,040 | 6.3% | 37,412 | 7,972 | 21.3% | - | - | |
| **Total** | <12 months | 72,601 | 9,691 | 13.3% | 35,026 | 8,091 | 23.1% | 4,094 | 1,761 | 43.0% |
| | 12 - 23 months | 37,406 | 10,477 | 28.0% | 28,885 | 9,584 | 33.2% | 3,908 | 1,994 | 51.0% |
| | 24 - 59 months | 14,154 | 2,648 | 18.7% | 6,476 | 2,013 | 31.1% | 3,323 | 1,337 | 40.2% |
| | < 5 yrs | 220,618 | 28,566 | 12.9% | 74,322 | 20,472 | 27.5% | 11,325 | 5,092 | 45.0% |
| | >=5 yrs | 24,708 | 754 | 3.1% | 2,624 | 184 | 7.0% | - | - | |
| | Total | 401,613 | 35,459 | 8.8% | 83,604 | 21,132 | 25.3% | - | - | |

#### Reporting checklists:

| Table S2: Guidelines for Accurate and Transparent Health Estimates Reporting (GATHER) Checklist (Stevens et al., 2016) | | |
|---|---|---|
| **Item #** | **Checklist item** | **Reported on page #** |
| **Objectives and funding** | | |
| **1** | Define the indicator(s), populations (including age, sex, and geographic entities), and time period(s) for which estimates were made. | Introduction paragraph 2 Materials and Methods paragraph 1 |
| **2** | List the funding sources for the work. | Abstract, Declarations, role of the funding source paragraph |
| **Data Inputs** | | |
| *For all data inputs from multiple sources that are synthesized as part of the study:* | | |
| **3** | Describe how the data were identified and how the data were accessed. | Methods, data sources paragraph |
| **4** | Specify the inclusion and exclusion criteria. Identify all ad-hoc exclusions. | Methods, data sources paragraph |
| **5** | Provide information on all included data sources and their main characteristics. For each data source used, report reference information or contact name/institution, population represented, data collection method, year(s) of data collection, sex and age range, diagnostic criteria or measurement method, and sample size, as relevant. | Methods, data sources paragraph |
| **6** | Identify and describe any categories of input data that have potentially important biases (e.g., based on characteristics listed in item 5). | Methods, Imputation paragraph |
| *For data inputs that contribute to the analysis but were not synthesized as part of the study:* | | |
| **7** | Describe and give sources for any other data inputs. | Methods, Imputation paragraph |
| *For all data inputs:* | | |
| **8** | Provide all data inputs in a file format from which data can be efficiently extracted (e.g., a spreadsheet rather than a PDF), including all relevant meta-data listed in item 5. For any data inputs that cannot be shared because of ethical or legal reasons, such as third-party ownership, provide a contact name or the name of the institution that retains the right to the data. | Supplementary file 3 |
| **Data analysis** | | |
| **9** | Provide a conceptual overview of the data analysis method. A diagram may be helpful. | Methods Figure 1 |

| Table S2: Guidelines for Accurate and Transparent Health Estimates Reporting (GATHER) Checklist (Stevens et al., 2016) | | |
|---|---|---|
| **Item #** | **Checklist item** | **Reported on page #** |
| **10** | Provide a detailed description of all steps of the analysis, including mathematical formulae. This description should cover, as relevant, data cleaning, data pre-processing, data adjustments and weighting of data sources, and mathematical or statistical model(s). | Methods, Statistical methods paragraph, Supplementary file 2 |
| **11** | Describe how candidate models were evaluated and how the final model(s) were selected. | Methods, Statistical methods paragraph |
| **12** | Provide the results of an evaluation of model performance, if done, as well as the results of any relevant sensitivity analysis. | Supplementary |
| **13** | Describe methods for calculating uncertainty of the estimates. State which sources of uncertainty were, and were not, accounted for in the uncertainty | Methods, Statistical methods paragraph |
| **14** | State how analytic or statistical source code used to generate estimates can be accessed. | Methods, data sources and Statistical methods paragraph |
| **Results and Discussion** | | |
| **15** | Provide published estimates in a file format from which data can be efficiently extracted. | Supplementary RIS files 1a and 1b |
| **16** | Report a quantitative measure of the uncertainty of the estimates (e.g. uncertainty intervals). | Throughout. Results, Tables, and Figures (and their data tables) |
| **17** | Interpret results in light of existing evidence. If updating a previous set of estimates, describe the reasons for changes in estimates. | Discussion |
| **18** | Discuss limitations of the estimates. Include a discussion of any modelling assumptions or data limitations that affect interpretation of the estimates. | Discussion |

| **Table S3: Standardized Reporting of Burden of Disease Studies (STROBOD) Checklist (Devleesschauwer et al., 2024)** | | |
|---|---|---|
| **Item #** | **Domains and description of the recommended items** | **Reported on page #** |
| **Title** | | |
| 1 | Identify the study as a burden of disease assessment by including keywords (e.g., Years of Life Lost, Years Lost due to Disability, Disability-Adjusted Life Years, burden of disease etc.), and describe the study setting | Title |
| **Abstract** | | |
| 2 | Provide a summary of objectives, study setting, methods (including data sources and key methodological design choices used), results (including point estimates and, if applicable, uncertainty intervals), and conclusions | Abstract |
| **Introduction** | | |
| 3 | Present background information to the study, its study aim(s), and its relevance for health policy or practice | Introduction |
| **Methods** | | |
| *Study Setting* | | |
| 4 | Report for which cause(s) the burden was calculated. Provide a case definition, e.g., in terms of an internationally recognized classification system such as the International Classification of Diseases and Related Health Problems 10th Revision | Introduction paragraph 1 |
| 5 | Report the reference population and any stratification of the reference population for the burden of disease assessment, i.e., the population for which the burden was calculated. This may include the geographical location (e.g., country or province/state), and whether the general population or a specific subset of the population (e.g., females, adolescents aged 10–19 years, etc.) was considered | Methods paragraph 2 |
| 6 | Report the reference time period (e.g., year(s), month(s)) of the study. This refers to the time period to which the burden of disease estimates refers. | Methods paragraph 1 |
| *Epidemiological and demographic input data* | | |
| 7 | Report the sources, values, ranges, and, if used, probability distributions for all epidemiological input parameters. Report reasons or sources for distributions used to represent uncertainty where appropriate. Providing a (supplementary) table to show all epidemiological input parameters and respective sources and assumptions is strongly recommended | Methods,<br>Approach 1<br>Approach 2 |
| 8 | Describe all possible data manipulations, such as bias corrections, data integration steps, or methods to ensure internal consistency of the data inputs | Methods, Imputation paragraph |

| Table S3: Standardized Reporting of Burden of Disease Studies (STROBOD) Checklist (Devleesschauwer et al., 2024) | | |
|---|---|---|
| **Item #** | **Domains and description of the recommended items** | **Reported on page #** |
| 9 | Report the sources and values of any population data used. If applicable, report the standard population used to calculate age-standardized rates | Methods, Approach 1: paragraph 1 Approach 2: paragraph 1 |
| *DALY methods* | | |
| 10 | Report the age-conditional life expectancy used for calculating Years of Life Lost (i.e., national, regional, or aspirational life tables) or other methods (e.g., potential years of life lost, proportion of premature deaths under a selected age threshold etc.) | Not applicable |
| 11 | Report the perspective taken for calculating Years Lost due to Disability, i.e., incidence or prevalence perspective | Not applicable |
| *Disease model* | | |
| 12 | Describe the disease model. Present and justify the included health outcomes and health states. Providing a (supplementary) figure visualizing the disease model is strongly recommended | Methods, Statistical methods paragraph, Supplementary file 2 |
| 13 | Report the source(s) and values of the used disability weights. Providing a (supplementary) table depicting the health states, brief lay descriptions, and the numerical values followed by its uncertainty intervals is strongly recommended | Reference 18 |
| 14 | If new disability weights were elicited, provide information on how the health states were described and the elicitation procedures. As a minimum to the latter, describe which valuation technique was used and which reference group and size of the group (also known as panel of judges) evaluated the health states. Providing a supplementary table with a description of the valuation technique and brief lay descriptions used is strongly recommended | Not applicable |
| 15 | Report the source(s) and values of the used durations (if applicable). Providing a (supplementary) table depicting the health states and the numerical values followed by its uncertainty intervals is strongly recommended | Not applicable |
| 16 | Report the source(s) and values of the used conditional probabilities, severity distribution, and/or transition rates. Providing a (supplementary) table depicting the parent/child health outcomes and health states and the numerical values followed by its uncertainty intervals is strongly recommended | Method, Data sources paragraph 3 |
| *Multimorbidity adjustments* | | |
| 17 | Report whether or not multimorbidity adjustments were applied to any of the input variables in the estimation of Years Lost due to | Not applicable |

| Table S3: Standardized Reporting of Burden of Disease Studies (STROBOD) Checklist (Devleesschauwer et al., 2024) | | |
|---|---|---|
| **Item #** | **Domains and description of the recommended items** | **Reported on page #** |
| | Disability. If applied, describe which multimorbidity adjustment method was used | |
| *Social weighting factors* | | |
| 18 | Report whether or not age weighting was applied. If applied, describe which parameters were used | Not applicable |
| 19 | Report whether or not time discounting was applied. If applied, describe which discount rate was used | Not applicable |
| *Uncertainty and scenario analysis* | | |
| 20 | Describe any methods used to perform uncertainty and variable importance (sensitivity) analyses. If, for example, Monte Carlo simulations were used, report the number of iterations | Method, Statistical method |
| 21 | Describe any scenario analyses that were performed. Present the rationale and the alternative data inputs defining the alternative scenarios | Not applicable |
| **Results** | | |
| 22 | Report the point estimates and, if applicable, the uncertainty interval of the burden of disease estimates. Provide both absolute values, crude rates (optional), and age- standardized rates per 100,000 in a table or figure | Results and Tables |
| 23 | If applicable, report the results of the scenario analyses. Tables and/or figures illustrating findings on the scenario analyses are strongly recommended | Results, figure 5 |
| **Discussion** | | |
| 24 | Summarize the key study findings and describe how they support the conclusions reached | Discussion |
| 25 | Discuss how the findings fit within current knowledge. Discuss potential implications for public health practice. Compare the results with those of other studies, and discuss methodological design differences, if relevant | Discussion |
| 26 | Discuss strengths and limitations, and the generalizability of the study findings. If applicable, discuss the results of the uncertainty and scenario analyses | Discussion |
| **Open science** | | |
| 27 | Make the source code or computational model(s) available as supporting information or via a dedicated open access repository (e.g., GitHub) | Data availability section |
| 28 | Describe how the study was funded and the role of the funder in the identification, design, conduct, and reporting of the analysis. Describe | Abstract, Declarations, role of |

| Table S3: Standardized Reporting of Burden of Disease Studies (STROBOD) Checklist (Devleesschauwer et al., 2024) | | |
|---|---|---|
| **Item #** | **Domains and description of the recommended items** | **Reported on page #** |
| | other non-monetary sources of support or any potential conflict(s) of interest of the study contributor(s) in accordance with the journal policy | the funding source paragraph |

| Table S4: Preferred Reporting Items for Systematic Reviews and Meta-Analyses (PRISMA) checklist (Page et al., 2021) | | | |
|---|---|---|---|
| **Section and Topic** | **Item #** | **Checklist item** | **Location where item is reported** |
| **TITLE** | | | |
| Title | 1 | Identify the report as a systematic review. | Title |
| **ABSTRACT** | | | |
| Abstract | 2 | See the PRISMA 2020 for Abstracts checklist. | Done |
| **INTRODUCTION** | | | |
| Rationale | 3 | Describe the rationale for the review in the context of existing knowledge. | Page 6 |
| Objectives | 4 | Provide an explicit statement of the objective(s) or question(s) the review addresses. | P7 |
| **METHODS** | | | |
| Eligibility criteria | 5 | Specify the inclusion and exclusion criteria for the review and how studies were grouped for the syntheses. | P8 |
| Information sources | 6 | Specify all databases, registers, websites, organisations, reference lists and other sources searched or consulted to identify studies. Specify the date when each source was last searched or consulted. | P8 and P12 |
| Search strategy | 7 | Present the full search strategies for all databases, registers and websites, including any filters and limits used. | Supplementary file 1 |
| Selection process | 8 | Specify the methods used to decide whether a study met the inclusion criteria of the review, including how many reviewers screened each record and each report retrieved, whether they worked independently, and if applicable, details of automation tools used in the process. | P8 |
| Data collection process | 9 | Specify the methods used to collect data from reports, including how many reviewers collected data from each report, whether they worked independently, any processes for obtaining or confirming data from study investigators, and if applicable, details of automation tools used in the process. | P8-9 |
| Data items | 10a | List and define all outcomes for which data were sought. Specify whether all results that were compatible with each outcome domain in each study were sought (e.g. for all measures, time points, analyses), and if not, the methods used to decide which results to collect. | P7 |
| | 10b | List and define all other variables for which data were sought (e.g. participant and intervention characteristics, funding sources). Describe any assumptions made about any missing or unclear information. | P7 |
| Study risk of bias assessment | 11 | Specify the methods used to assess risk of bias in the included studies, including details of the tool(s) used, how many reviewers assessed each study and whether they worked independently, and if applicable, details of automation tools used in the process. | P7 |
| Effect measures | 12 | Specify for each outcome the effect measure(s) (e.g. risk ratio, mean difference) used in the synthesis or presentation of results. | P10 and P13 |
| Synthesis | 13a | Describe the processes used to decide which studies were eligible for each synthesis (e.g. tabulating the | P8 |

**Table S4: Preferred Reporting Items for Systematic Reviews and Meta-Analyses (PRISMA) checklist (Page et al., 2021)**

| Section and Topic | Item # | Checklist item | Location where item is reported |
|---|---|---|---|
| methods | | study intervention characteristics and comparing against the planned groups for each synthesis (item #5)). | |
| | 13b | Describe any methods required to prepare the data for presentation or synthesis, such as handling of missing summary statistics, or data conversions. | P12 |
| | 13c | Describe any methods used to tabulate or visually display results of individual studies and syntheses. | P10 and P13 |
| | 13d | Describe any methods used to synthesize results and provide a rationale for the choice(s). If meta-analysis was performed, describe the model(s), method(s) to identify the presence and extent of statistical heterogeneity, and software package(s) used. | P10 and P13 |
| | 13e | Describe any methods used to explore possible causes of heterogeneity among study results (e.g. subgroup analysis, meta-regression). | P10 and P13 |
| | 13f | Describe any sensitivity analyses conducted to assess robustness of the synthesized results. | Supplementary |
| Reporting bias assessment | 14 | Describe any methods used to assess risk of bias due to missing results in a synthesis (arising from reporting biases). | P11 and P14 |
| Certainty assessment | 15 | Describe any methods used to assess certainty (or confidence) in the body of evidence for an outcome. | P11 and P14 |
| **RESULTS** | | | |
| Study selection | 16a | Describe the results of the search and selection process, from the number of records identified in the search to the number of studies included in the review, ideally using a flow diagram. | P14, Figure 1 |
| | 16b | Cite studies that might appear to meet the inclusion criteria, but which were excluded, and explain why they were excluded. | Figure 1 |
| Study characteristics | 17 | Cite each included study and present its characteristics. | Figure 1 |
| Risk of bias in studies | 18 | Present assessments of risk of bias for each included study. | Not done |
| Results of individual studies | 19 | For all outcomes, present, for each study: (a) summary statistics for each group (where appropriate) and (b) an effect estimate and its precision (e.g. confidence/credible interval), ideally using structured tables or plots. | Table 1 |
| Results of syntheses | 20a | For each synthesis, briefly summarise the characteristics and risk of bias among contributing studies. | Not done |
| | 20b | Present results of all statistical syntheses conducted. If meta-analysis was done, present for each the summary estimate and its precision (e.g. confidence/credible interval) and measures of statistical heterogeneity. If comparing groups, describe the direction of the effect. | P16-17 |
| | 20c | Present results of all investigations of possible causes of heterogeneity among study results. | Table 1-3 |
| | 20d | Present results of all sensitivity analyses conducted to assess the robustness of the synthesized results. | Supplementary |

**Table S4: Preferred Reporting Items for Systematic Reviews and Meta-Analyses (PRISMA) checklist (Page et al., 2021)**

| Section and Topic | Item # | Checklist item | Location where item is reported |
|---|---|---|---|
| Reporting biases | 21 | Present assessments of risk of bias due to missing results (arising from reporting biases) for each synthesis assessed. | |
| Certainty of evidence | 22 | Present assessments of certainty (or confidence) in the body of evidence for each outcome assessed. | |
| **DISCUSSION** | | | |
| Discussion | 23a | Provide a general interpretation of the results in the context of other evidence. | P21-27 |
| | 23b | Discuss any limitations of the evidence included in the review. | P28-29 |
| | 23c | Discuss any limitations of the review processes used. | P28-29 |
| | 23d | Discuss implications of the results for practice, policy, and future research. | P29-30 |
| **OTHER INFORMATION** | | | |
| Registration and protocol | 24a | Provide registration information for the review, including register name and registration number, or state that the review was not registered. | Not done |
| | 24b | Indicate where the review protocol can be accessed, or state that a protocol was not prepared. | Not done |
| | 24c | Describe and explain any amendments to information provided at registration or in the protocol. | Not done |
| Support | 25 | Describe sources of financial or non-financial support for the review, and the role of the funders or sponsors in the review. | WHO, NIH (no role) |
| Competing interests | 26 | Declare any competing interests of review authors. | None |
| Availability of data, code and other materials | 27 | Report which of the following are publicly available and where they can be found: template data collection forms; data extracted from included studies; data used for all analyses; analytic code; any other materials used in the review. | P33 |

*From:* Page MJ, McKenzie JE, Bossuyt PM, Boutron I, Hoffmann TC, Mulrow CD, et al. The PRISMA 2020 statement: an updated guideline for reporting systematic reviews. BMJ 2021;372:n71. doi: 10.1136/bmj.n71. This work is licensed under CC BY 4.0. To view a copy of this license, visit https://creativecommons.org/licenses/by/4.0/
